# High-dimensional analysis reveals abnormal B cell subsets associated with specific changes to circulating T and myeloid cell populations in patients with idiopathic inflammatory myopathies

**DOI:** 10.1101/2021.03.23.21253635

**Authors:** Erin M. Wilfong, Todd Bartkowiak, Katherine N. Vowell, Camille S. Westlake, Jonathan M. Irish, Peggy L. Kendall, Leslie J. Crofford, Rachel H. Bonami

## Abstract

**Objectives:** The idiopathic inflammatory myopathies (IIM) are a clinically heterogeneous group of conditions affecting the skin, muscle, joint, and lung in various combinations. This study aims to investigate the immunologic heterogeneity through detailed immunophenotyping of peripheral blood mononuclear cells (PBMCs) in IIM patients and healthy controls.

**Methods:** We collected PBMCs from 17 patients with a clinical diagnosis of inflammatory myositis in the inpatient or outpatient setting and performed immunophenotyping using mass cytometry by time of flight (CyTOF) to simultaneously characterize B, T, and myeloid cell subsets. Data were analyzed using a combination of supervised biaxial gating and unsupervised clustering algorithms including t-distributed stochastic neighbor embedding (tSNE), cluster identification, characterization, and regression (CITRUS), and marker enrichment modeling (MEM).

**Results:** We identified two distinct immune signatures amongst IIM patients. In one signature, increased CD19+CXCR4hiCCR7hi cells correlated with increased CD3+CXCR4hiCD38hi (r=0.62, p=0.009) and CD14+CD16-CXCR4+CD38+HLADR-(r=0.61, p=0.01) populations. In the second signature, increased CD19+CD21loCD11c+ cells correlated with an increased CD3+CD4+PD1+ (r=0.60, p=0.01) population. Other shared immunologic features amongst IIM patients compared to healthy controls included decreased surface expression of RP105/CD180 on B cells (median mass intensity 39.9 ± 16.0 v. 60.9 ± 20.1, p=0.002). In the T cell compartment, all circulating CD3+CXCR3+ subsets (2.7 ± 2.4 v. 9.6 ± 8.1% of all PBMCs, p=0.0004) were reduced.

**Conclusion:** Based on circulating B cell phenotype, we identified two distinct immunologic signatures in IIM patients. Future work is needed to determine the significance of these immune signatures for clinical manifestations and treatment responses.

## Introduction

A major challenge in diagnosing and managing the idiopathic inflammatory myopathies (IIM) is vast clinical heterogeneity. While initial criteria focused on proximal muscle weakness and classic skin rashes [1], there is increasing recognition that patients present with varying cutaneous, muscular, and pulmonary involvement. Indeed, perhaps the best predictor of disease manifestations is the autoantibody identified for a given patient. Non-Jo-1-related anti-tRNA synthetase syndrome (ARS) patients frequently present with prominent interstitial lung disease (ILD) whereas Jo-1-related ARS patients have classical dermatomyositis [2]. Mi-2 positive patients frequently manifest with classical rashes, mild to moderate muscle involvement, and rarely have ILD [3]. Given the role of autoantibodies in IIM and the efficacy of rituximab in treating IIM [4], particularly amongst seropositive IIM patients [5], it is likely that B cells play a critical role in disease pathogenesis and progression.

Beyond secreting potentially pathogenic antibodies [6], B cells also express these antibodies on the cell surface as the B cell receptor (BCR). After recognizing cognate antigen using the BCR, B cells internalize, process, and present cognate antigen via major histocompatibility complex class II (MHCII) molecules to activate CD4+ T cells [7]. This ability to activate T cells is critically involved in the development of autoimmune disease, and B cells are required for murine models of spontaneous autoimmune thyroiditis [8], type 1 diabetes mellitus [9, 10], Sjögren’s syndrome [11], and systemic lupus erythematosus [12]. Beyond activation of CD4+ T cells, B cells are also capable of polarizing other T cells [13-15] and myeloid cells [16, 17].

A key limitation in prior IIM immunophenotyping studies [18-20] was the number of parameters that could be simultaneously studied using fluorescence cytometry. While detailed B, T, or myeloid cell phenotyping could be performed individually, the number of parameters required to simultaneously evaluate all PBMC subsets was not feasible. Mass <u>cy</u>tometry by time <u>o</u>f flight (CyTOF) uses isotopically pure heavy metals conjugated to antibodies for target detection [21]; at present, 45 antibody markers can be detected via mass cytometry compared to 28 markers by spectral fluorescence flow cytometry [22]. Further, multiple unsupervised analytical tools (e.g. t-distributed stochastic neighbor embedding (tSNE) [23], cluster identification, characterization, and regression (CITRUS) [24], and Flow Self Organized Maps (FlowSOM) [25]), help identify previously unrecognized immune populations that express unexpected combinations of phenotypic markers. Here, we define immunophenotypic differences between IIM patients compared to healthy controls identified by CyTOF using both supervised (biaxial gating) and unsupervised (tSNE, CITRUS) analytic techniques.

## Methods

### Patients

Institutional Review Board approval was obtained (VUMC IRB 141415). Patients clinically diagnosed with IIM were enrolled into the Myositis and Scleroderma Treatment Initiative Center (MYSTIC) Cohort in either the outpatient clinic or inpatient setting at Vanderbilt University Medical Center between 9/17/2017 and 9/30/2018. Individuals enrolling as healthy controls completed a health questionnaire to verify a negative review of systems and no personal or family history of autoimmunity in a first degree relative. We performed clinical phenotyping by chart abstraction to estimate the date of symptom onset and collected serologic data, including anti-nuclear antibodies (ANA), rheumatoid factor (RF), cyclic citrullinated peptide (CCP), and an extended myositis panel obtained through ARUP (Salt Lake City, UT). We defined patients as having ILD if a radiologist determined that fibrosis was present on a CT scan. If the treating clinician escalated immunosuppression, the patient was defined as having active disease. Clinical data is reported as the mean ± standard deviation unless otherwise indicated. We isolated peripheral blood mononuclear cells (PBMCs) from blood collected in sodium heparin CPT tubes (BD Biosciences, San Jose CA) per manufacturer’s directions and cryopreserved for future study.

### Mass cytometry

Seventeen IIM patients with active disease and eighteen healthy controls were included for CyTOF analysis. For CyTOF acquisition, we thawed 3-5 million PBMCs per individual, viability stained with cisplatin, and stained for surface and intracellular markers (See supplemental methods and tables S1). Data were acquired using a CyTOF Helios 3.0 (Fluidigm Sciences, Sunnyvale, CA) and CyTOF software (version 6.7.1014) at the Vanderbilt University Medical Center Mass Cytometry Center of Excellence. Dual count calibration and noise reduction were applied prior to acquisition; 100,000-400,000 events were collected per sample.

### Flow cytometry

We selected six representative IIM patients and 6 healthy controls for flow cytometric studies of CD180 expression, for which 4-5 million cells per individual were thawed and stained (see supplemental methods and table S2). All data were acquired on a BD LSRII Fortessa instrument.

### Data analysis

Mass cytometry FCS files underwent Fluidigm bead normalization and analysis using Cytobank software per established methods [26]. Supervised (traditional biaxial gating) or semi-supervised (tSNE [23] or CITRUS [24]) analyses were conducted using CytoBank (Santa Clara, CA). Marker enrichment modeling labels [27] aided in determining biaxial gating schemes for CITRUS-identified populations. Details of mass cytometric data analysis are included in the supplemental methods and tables S4-S5. Fluorescence cytometry data was analyzed using FlowJo version 9.9.6.

### Statistical methods

Population statistics were exported from CytoBank or FlowJo as appropriate and analyzed with Prism software (GraphPad, La Jolla, CA) to calculate descriptive statistics. If multiple groups were compared, we performed a Kruskal-Wallis ANOVA and, if p<0.05, post-hoc Mann-Whitney U-tests were performed. For comparison of two continuous variables, we utilized Mann-Whitney U-tests. For comparison of two dichotomous variables, a Fisher’s exact test was performed. Spearman’s correlation coefficients were utilized to identify the presence of a statistical correlation between populations.

### Patient and Public Involvement Statement

While patients did not participate in study design, patients donated blood to the MYSTIC cohort knowing of our plans to perform immunophenotyping by mass cytometry at the time of enrollment through study brochures. Aggregated, but not individual, data will be available to participating patients.

## Results

### Patient Characteristics

We studied seventeen patients clinically diagnosed with IIM and 18 healthy controls. Basic demographics and clinical information are shown in Table 1. Detailed clinical phenotyping is shown in Table S5. IIM patients were slightly older than healthy controls (56.8 ± 12.0 v. 46.4 ± 12.0 years, p=0.01). Seven patients were receiving corticosteroids at the time of enrollment; one patient was taking methotrexate at the time of enrollment with active skin, lung, and joint involvement. Eleven patients met the 2017 classification criteria for probable or definite IIM. Those not meeting classification criteria all had myositis-specific autoantibodies.

**Table 1.**
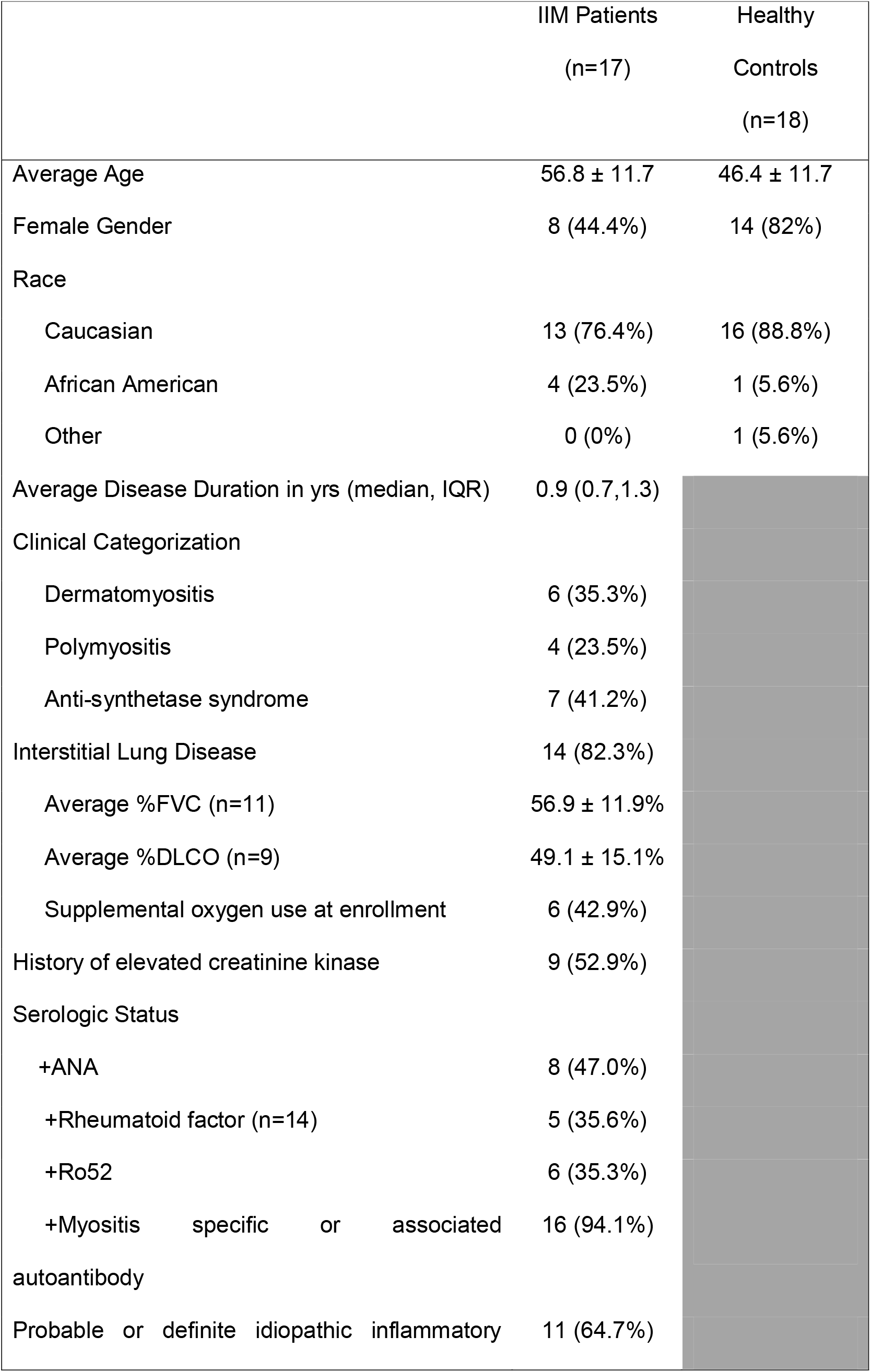
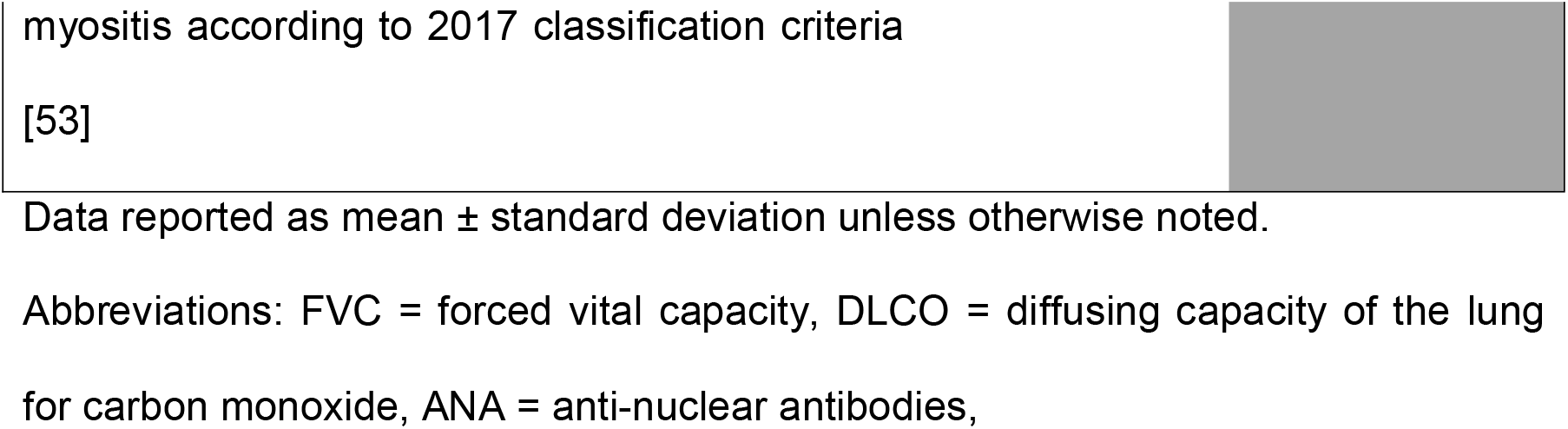
Patient Demographics

|  | IIM Patients<br>(n=17) | Healthy<br>Controls<br>(n=18) |
| --- | --- | --- |
| Average Age | 56.8 ± 11.7 | 46.4 ± 11.7 |
| Female Gender | 8 (44.4%) | 14 (82%) |
| Race |  |  |
| Caucasian | 13 (76.4%) | 16 (88.8%) |
| African American | 4 (23.5%) | 1 (5.6%) |
| Other | 0 (0%) | 1 (5.6%) |
| Average Disease Duration in yrs (median, IQR) | 0.9 (0.7,1.3) |  |
| Clinical Categorization |  |  |
| Dermatomyositis | 6 (35.3%) |  |
| Polymyositis | 4 (23.5%) |  |
| Anti-synthetase syndrome | 7 (41.2%) |  |
| Interstitial Lung Disease | 14 (82.3%) |  |
| Average %FVC (n=11) | 56.9 ± 11.9% |  |
| Average %DLCO (n=9) | 49.1 ± 15.1% |  |
| Supplemental oxygen use at enrollment | 6 (42.9%) |  |
| History of elevated creatinine kinase | 9 (52.9%) |  |
| Serologic Status |  |  |
| +ANA | 8 (47.0%) |  |
| +Rheumatoid factor (n=14) | 5 (35.6%) |  |
| +Ro52 | 6 (35.3%) |  |
| +Myositis specific or associated autoantibody | 16 (94.1%) |  |
| Probable or definite idiopathic inflammatory | 11 (64.7%) |  |

Data reported as mean ± standard deviation unless otherwise noted.
Abbreviations: FVC = forced vital capacity, DLCO = diffusing capacity of the lung for carbon monoxide, ANA = anti-nuclear antibodies,

### Lymphoid and myeloid subsets are altered in IIM patients compared to healthy controls

We evaluated common myeloid, T, and B cell subsets using tSNE (Figure 1A-B) and traditional biaxial gating of CyTOF data (Supplemental Figure S1 and Figure 1E) based on standardized immunophenotype markers [28]. Table 2 shows the average, standard deviation, and p value comparing all populations between IIM and healthy controls. Complete blood count with differential was available for 14 IIM patients and 14 healthy controls. There was no difference between the number of circulating PBMCs between IIM patients and healthy controls (Figure 1D). As shown in Fig. 1E, IIM patients had a decreased frequency of class-switched (CD19+CD27+IgM-) and non-class-switched (CD19+CD27+IgM+) memory B cells compared to healthy controls; there was no difference in the frequency of naïve (CD19+CD27-IgD+) or total B cells. While there was no statistically significant difference in CD8+ T cells, there was a decrease in CD4+, CD4-CD8- and CD4+CXCR5+PD1+ T cells. Classical monocytes (CD14+CD16-CD19-CD3-) were increased in IIM patients compared to healthy controls, but there was no difference in the frequency of natural killer cells (CD16+CD19-CD3-CD14-) or non-classical monocytes (CD14+CD16+CD19-CD3-).

**Table 2.** Peripheral Blood Mononuclear Cell Subsets and Median Mass Intensities (MMI) in Healthy Controls and IIM Patients

| Population | Healthy<br>(n=18) | IIM<br>(n=17) | P value |
| --- | --- | --- | --- |
| **Total number of circulating PBMCs (cells/mL)** | 2443 ± 614 | 2649 ± 1301 | 0.72 |
| **CD19+** |  |  |  |
| Total CD19<sup>+</sup> | 10.3 ± 4.3% | 10.8 ± 7.8 | 0.61 |
| CD19+CD27- (naïve)<sup>‡</sup> | 7.5 ± 4.0% | 8.6 ± 6.9% | 0.96 |
| CD19+CD27+IgM+ (non-class switched memory)<sup>‡</sup> | 0.9 ± 0.6% | 0.3 ± 0.3% | <0.0001 |
| CD19+CD27+IgM- (class switched memory)<sup>‡</sup> | 0.7 ± 0.3% | 0.4 ± 0.3% | 0.03 |
| CD19+CD24hiCD38hi<sup>‡</sup> | 3.8 ± 1.7% | 4.9 ± 5.1% | 0.66 |
| CD19+CD21lo<sup>‡</sup> | 5.8 ± 2.2% | 11.7 ± 7.2% | 0.001 |
| CD19+CD27-IgD- (DN B cells)<sup>‡</sup> | 7.4 ± 2.9% | 12.7 ± 5.8% | 0.002 |
| CD19+CD27-IgD+IgM- (BND cells)<sup>‡</sup> | 2.7 ± 1.0% | 2.6 ± 1.7% | 0.33 |
| CD19+ CD180 MMI | 60.9 ± 20.8 | 39.8 ± 16.0 | 0.002 |
| CD19+CD27- CD180 MMI | 58.3 ± 22.5 | 38.5 ± 16.3 | 0.009 |
| CD19+CD27+IgM+ CD180 MMI | 80.9 ± 19.1 | 58.5 ± 15.2 | 0.0005 |
| CD19+CD27+IgM- CD180 MMI | 73.5 ± 19.0 | 38.2 ± 28.5 | <0.0001 |
| CD19+CXCR4hiCCR7hiCD21lo (tSNE)<sup>‡</sup> | 0.1 ± 0.1% | 6.1 ± 19.1 % | 0.0001 |
| CD19+CD21loCD11c+ (tSNE)<sup>‡</sup> | 2.3 ± 1.2% | 4.1 ± 3.8% | 0.09 |
| **CD3+** |  |  |  |
| CD3+CXCR3<sup>+</sup> | 9.6 ± 8.1% | 2.6 ± 2.4% | 0.0004 |
| CD3+CXCR3<sup>-</sup> | 45.6 ± 9.8% | 37.1 ± 13.4% | 0.08 |
| **CD3+CD4+** |  |  |  |
| Total CD3+CD4<sup>+</sup> | 35.1 ± 6.6% | 24.3 ± 7.8% | 0.0001 |
| CD4+ Naive<sup>§</sup> | 27.7 ± 9.0% | 31.8 ± 17.8% | 0.89 |
| CD4+ Central Memory<sup>§</sup> | 18.5 ± 4.9% | 15.7 ± 5.9% | 0.23 |
| CD4+ Effector Memory<sup>§</sup> | 13.5 ± 3.9% | 11.3 ± 6.9% | 0.13 |
| CD4+ Effector<sup>§</sup> | 2.2 ± 1.2% | 3.4 ± 3.0% | 0.19 |
| Th0<sup>§</sup> | 27.9 ± 8.8% | 31.9 ± 16.1% | 0.64 |
| Th1<sup>§</sup> | 2.8 ± 2.0% | 0.8 ± 0.8% | 0.0002 |
| Th1Th17<sup>§</sup> | 1.1 ± 2.1% | 0.1 ± 0.1% | 0.01 |
| Th2<sup>§</sup> | 10.1 ± 4.6% | 11.1 ± 4.7% | 0.66 |
| Th2CXCR3+<sup>§</sup> | 1.9 ± 1.3% | 0.8 ± 0.6% | 0.003 |
| Th17<sup>§</sup> | 1.5 ± 1.4% | 1.3 ± 1.2% | 0.57 |
| Th17CXCR3+<sup>§</sup> | 0.6 ± 0.8% | 0.1 ± 0.1% | 0.03 |
| CD3+CD4+CXCR5+PD1+ (T follicular helper)<sup>§</sup> | 0.4 ± 0.2% | 0.1 ± 0.09% | 0.0001 |
| CD3+CD4+CCR4+CD25+CD127- (Treg)<sup>§</sup> | 2.2 ± 0.8 | 2.7 ± 2.0 | 0.88 |
| CD3+CD4+CXCR4hiCD38-(Cluster "A")<sup>§</sup> | 0.3 ± 0.3% | 5.5 ± 8.6 | <0.0001 |
| CD3+CD4+CD27-PD1+ (Cluster "B")<sup>§</sup> | 1.5 ± 1.6% | 3.2 ± 3.3% | 0.25 |
| **CD3+CD8+** |  |  |  |
| Total CD3+CD8+<sup>†</sup> | 13.7 ± 4.3% | 11.9 ± 8.1% | 0.40 |
| CD3+CD8+CXCR3+<sup>§</sup> | 7.6 ± 5.6% | 4.2 ± 4.6% | 0.03 |
| CD3+CD8+CXCR3-<sup>§</sup> | 16.1 ± 6.9% | 22.5 ± 11.5 | 0.06 |
| CD8+ Naïve<sup>§</sup> | 5.5 ± 2.3% | 4.5 ± 3.5% | 0.26 |
| CD8+ Central Memory<sup>§</sup> | 3.3 ± 1.3% | 1.8 ± 1.4% | 0.002 |
| CD8+ Effector Memory<sup>§</sup> | 8.9 ± 4.1% | 8.8 ± 8.3% | 0.42 |
| CD8+ Effector<sup>§</sup> | 2.6 ± 2.7% | 7.1 ± 6.8% | 0.02 |
| **CD3-CD4-CD8-** |  |  |  |
| Total CD3+CD4-CD8-<sup>†</sup> | 6.3 ± 4.7% | 3.2 ± 2.7% | 0.006 |
| CD3-CD4-CD8-CXCR3+<sup>§</sup> | 4.7 ± 6.5% | 1.0 ± 0.7% | 0.0002 |
| CD3-CD4-CD8-CXCR3-<sup>§</sup> | 5.7 ± 3.2% | 6.0 ± 4.0% | 0.71 |
| **CD19-CD3-** |  |  |  |
| CD3-CD19-CD14+ Classical Monocytes<sup>†</sup> | 5.3 ± 2.1% | 13.9 ± 9.1% | 0.0003 |
| CD3-CD19-CD14+CD16+ Non-classical monocytes<sup>†</sup> | 6.2 ± 1.8% | 8.6 ± 4.4% | 0.10 |
| CD3-CD19-CD14-CD16+ NK cells<sup>†</sup> | 9.2 ± 4.1% | 10.6 ± 6.3% | 0.66 |
| Biaxial "Cluster C"<sup>□</sup> | 2.3 ± 1.4% | 6.3 ± 5.3 % | 0.0029 |
| Biaxial "Cluster D"<sup>□</sup> | 1.9 ± 1.2% | 3.9 ± 2.0% | 0.0001 |

Values reported as average ± standard deviation. Statistical comparisons performed using Mann-Whitney U-test.
†frequency of all peripheral blood mononuclear cells, <sup>†</sup>frequency of all CD19+ cells, <sup>§</sup>frequency of all CD3+ cells, <sup>□</sup>frequency of all CD3-CD19- cells

**Figure 1.**
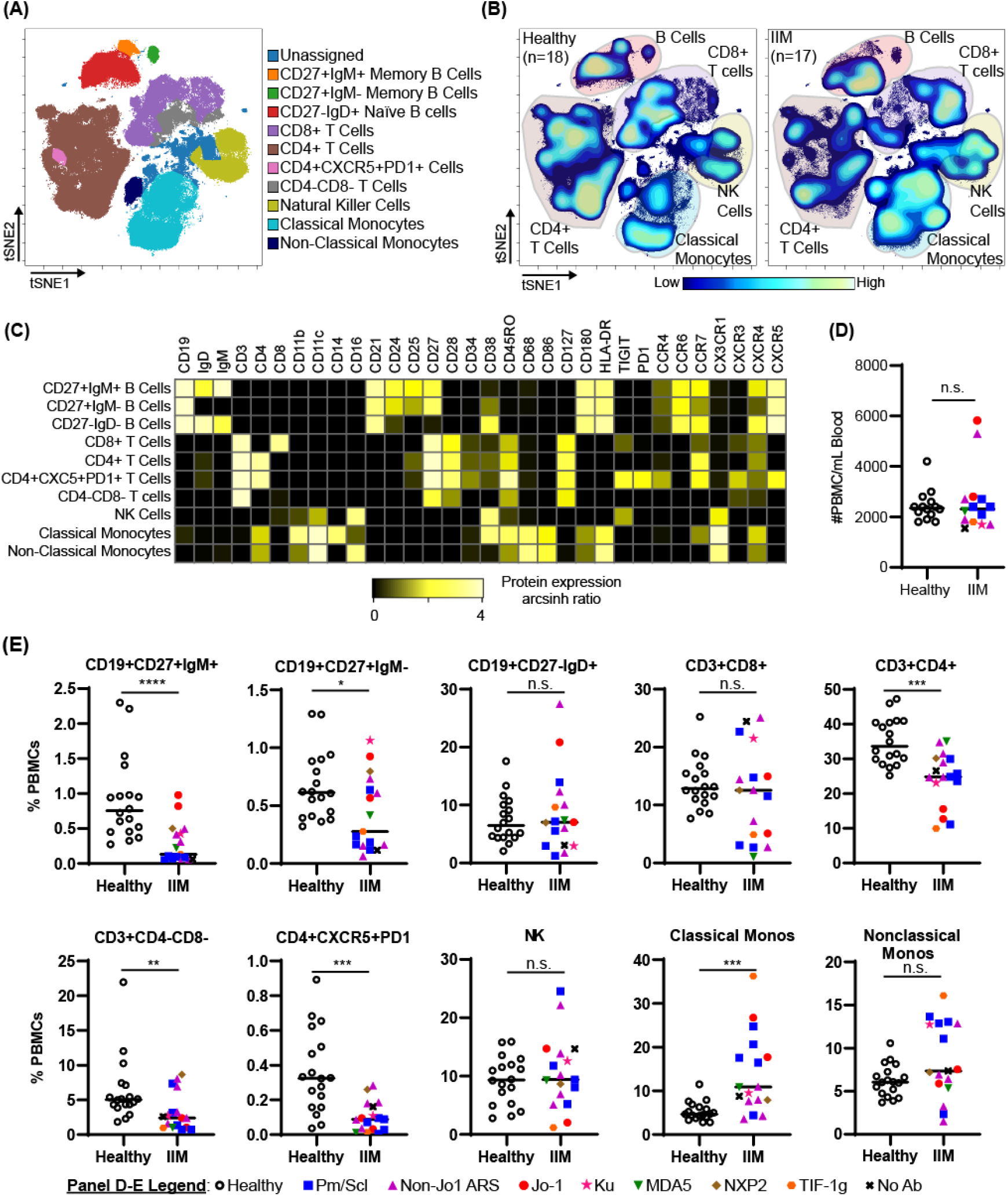
IIM patients show differences in peripheral blood immune subsets from healthy controls. (A) PBMCs from healthy controls (n=18) and IIM patients (n=17) were clustered using tSNE and pseudocolored to show major PBMC subsets. (B) Density contour maps of concatenated healthy controls versus IIM patients. (C) Expression heatmaps of representative markers for tSNE islands displaying the arcsinh ratio by table’s minimum. (D-E) Clinical seropositivity for the indicated autoantibodies is shown by legend symbols at the bottom of Panel E. Individual donors are plotted. (D) Absolute number of circulating PBMCs for healthy controls (n=14) and IIM patients (n=14) for whom count data were available. (E) Comparison of major circulating PBMC subsets between healthy controls and IIM patients using tSNE gating from panel A. Statistical comparisons were performed using Mann-Whitney U tests. *p<0.05, **p<0.01, ***p<0.001, ****p<0.0001.

### Autoimmune-prone CD21lo and DN B cell subsets are increased in IIM compared to healthy controls

Next, we investigated the previously described autoreactive CD24hiCD38hi transitional B cell [29], CD21lo/negative B cell [30], CD27-IgD-DN B cell [31], and B_ND_ cell [32] subsets as shown in Figure 2A. There was no difference in the frequency of CD24hiCD38hi transitional B cells or B_ND_ cells in IIM patients compared to healthy controls. However, IIM patients had increased CD21lo/neg and DN B cells compared to healthy controls (Figure 2B).

**Figure 2.**
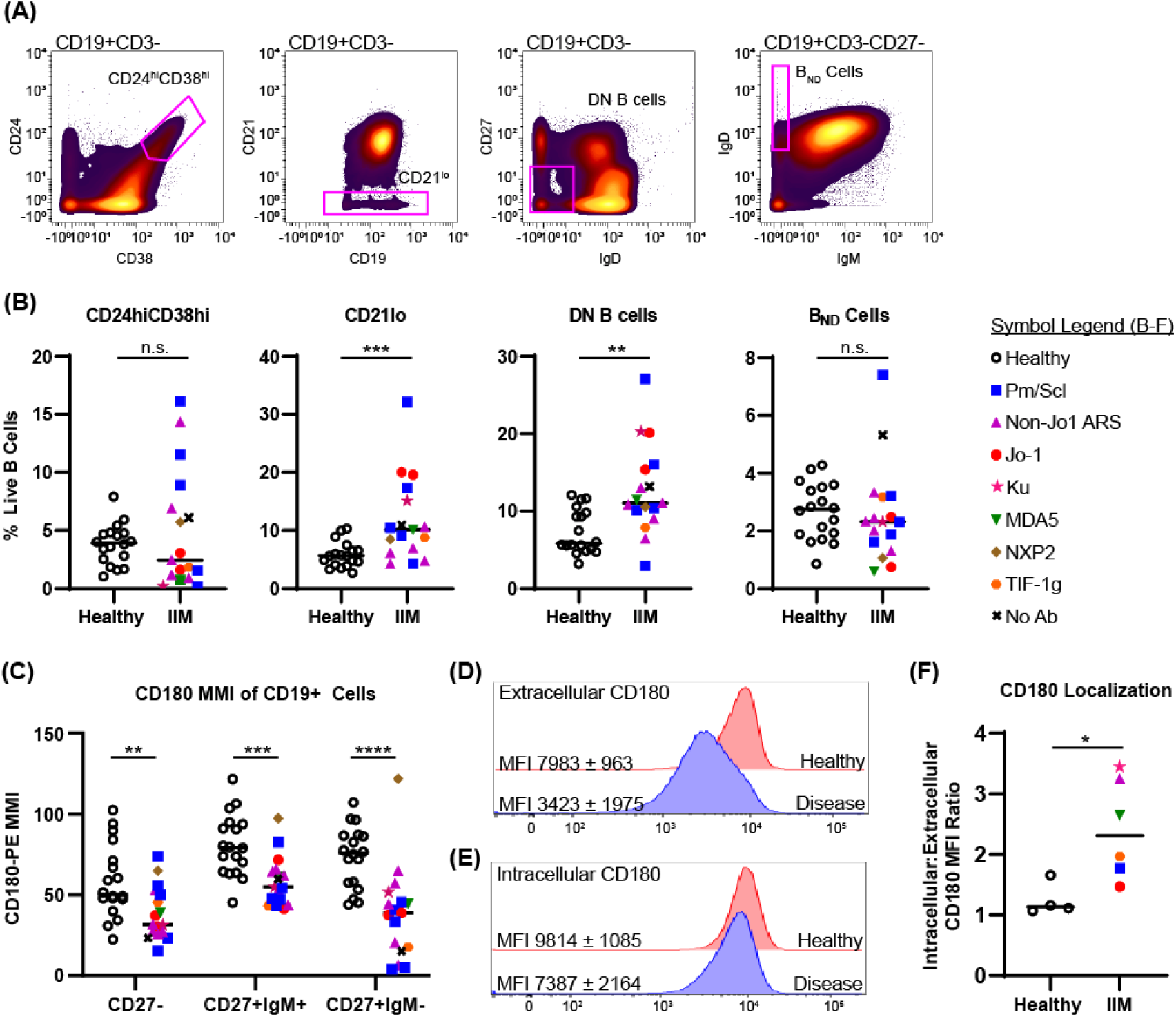
The frequency of autoreactive-prone CD21lo and DN B cells as well as surface expression of CD180 are reduced in IIM patients. (A) Mass cytometry gating scheme to identify CD19+CD24hiCD38hi transitional B cells, CD19+CD21lo cells, CD19+CD27-IgD-(DN B) cells, and CD27-IgD+IgM-(BND) cells in healthy controls (n=18) and IIM patients (n=17). (B) Population frequencies for panel A subsets are shown for each donor; symbols represent clinical autoantibody status as in Fig. 1. (C) CD180/RP105 mean mass intensity (MMI) for naive and memory CD19+ populations. (D) Fluorescent cytometry data quantifying CD180 expression on the cell surface and (E) intracellularly for four heathy controls and six IIM patients. (F) Ratio of intracellular to extracellular CD180 mean fluorescence intensity (MFI). Data are expressed as the mean ± standard deviation. Statistical comparisons were performed using Mann-Whitney U tests, *p<0.05, **p<0.01, ***p<0.001, ****p<0.0001.

Kikuchi et al. reported an RP105/CD180 lo B cell population that was increased in dermatomyositis [33]. Examination of CD180 median mass intensity (MMI) revealed decreased expression on naïve, class-switched memory, and non-class switched memory B cells in IIM patients compared to healthy donors (Figure 2C). To evaluate whether surface CD180 expression was decreased due to global protein downregulation, fluorescence cytometry was performed on 6 IIM patients and 4 healthy controls to quantify extracellular versus intracellular protein. Extracellular CD180 median fluorescent intensity was decreased in IIM patients compared to healthy controls, whereas intracellular CD180 expression was not different (Figure 2D-E).

### Circulating CXCR3+ T cells are decreased in IIM

We used previously defined biaxial gating schemes [28, 34] to evaluate CD4+ T cell subsets (Figure 3A and Supplemental Figure S2A). There were no differences in CD4+ naïve, central memory, effector memory, or effector cells between IIM patients and healthy controls (Supplemental Figure S2B). IIM patients had an increased frequency of CD8+ effector cells but not CD8+ naïve, central memory or effector memory cells (Supplemental Figure S2C). However, as shown in Figure 3C, all CXCR3+ Th subsets, including Th1, Th1Th17, CXCR3+Th2, and CXCR3+Th17 cells, were strikingly decreased in IIM compared to healthy controls. There was no difference in the CXCR3-subsets Th0, Th2, and Th17. We additionally determined that the CXCR3+ subsets of CD8+ and CD4-CD8-T cells were decreased in IIM compared to healthy controls, but there was again no difference in the CXCR3-subsets (Figure 3D-E).

**Figure 3.**
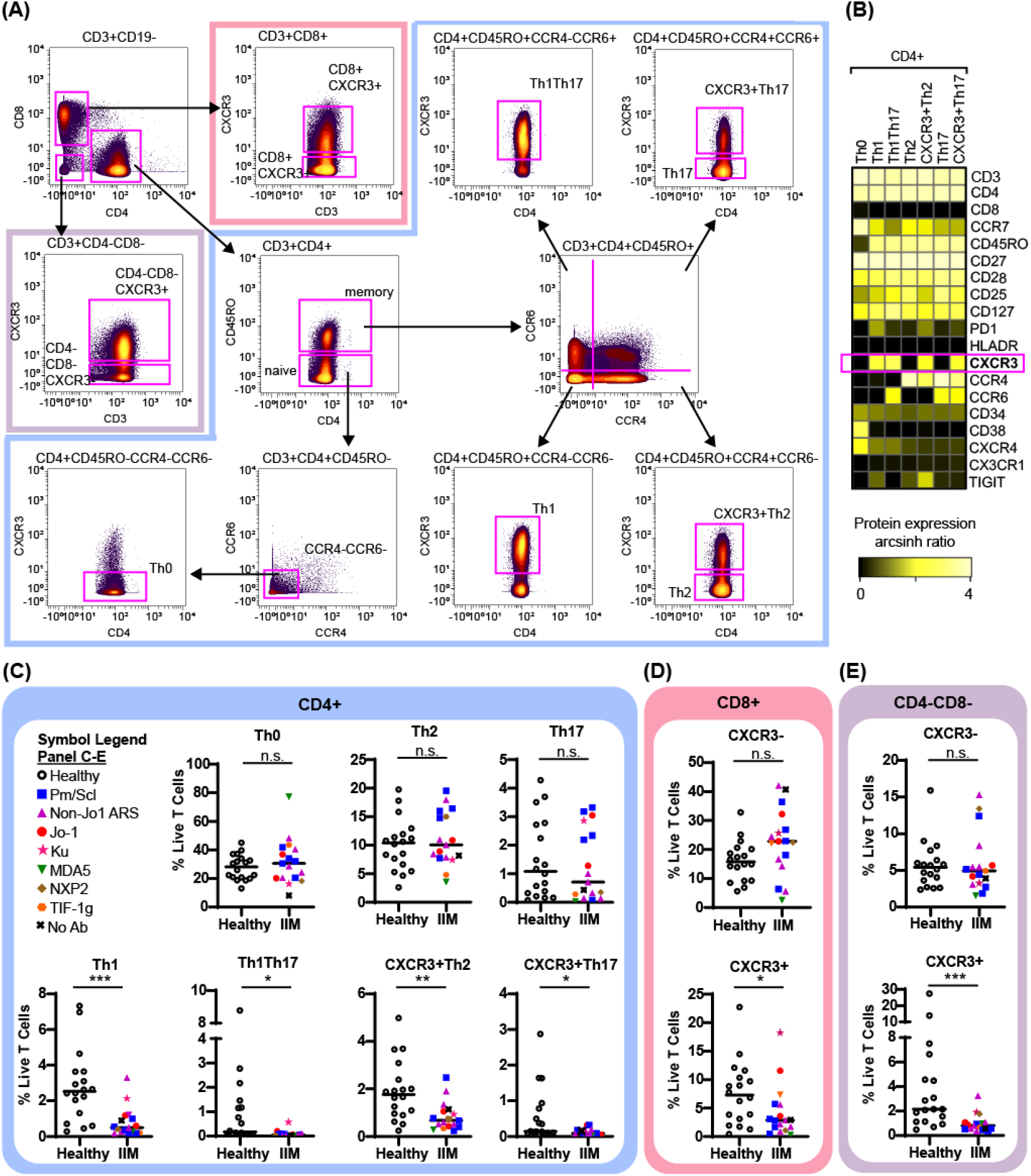
CXCR3+ T cell subsets are decreased in IIM. (A) Mass cytometry biaxial gating scheme to identify subsets of CD4-CD8-, CD4+, and CD8+ T cells for healthy controls (n=18) and IIM patients (n=17). (B) Expression heatmap of surface markers for CD4+ subsets displaying the arcsinh ratio by table’s minimum. (C-E) CXCR3- and CXCR3+ T cell populations were examined. Frequencies of biaxially gated T cell subsets are shown for each donor; clinical autoantibody status is indicated as in Fig. 1; (C) CD4+, (D) CD8+, and (E) CD4-CD8-T cell populations. Statistical comparisons were performed using Mann-Whitney U tests. *p<0.05, **p<0.01, ***p<0.001, ****p<0.0001.

### Unsupervised supervised analysis reveals two different abnormal CD19+ cell populations in IIM

Population identification using biaxial gating strategies relies on prior phenotypic marker knowledge. However, tSNE plots represent multidimensional cellular information in 2D space allowing for unbiased population identification and visual comparison [23]. The tSNE plots in Figure 4A show two abnormal B cell subsets with varying frequencies in IIM patients as well as the previously seen decrease in class-switched and non-class switched memory B cell subsets (Figure 4C). The CD19+CXCR4hiCCR7hi subset was increased in IIM patients compared to healthy controls. The second CD19+CD21loCD11c+ subset did not reach statistical significance but was increased in some IIM patients.

**Figure 4.**
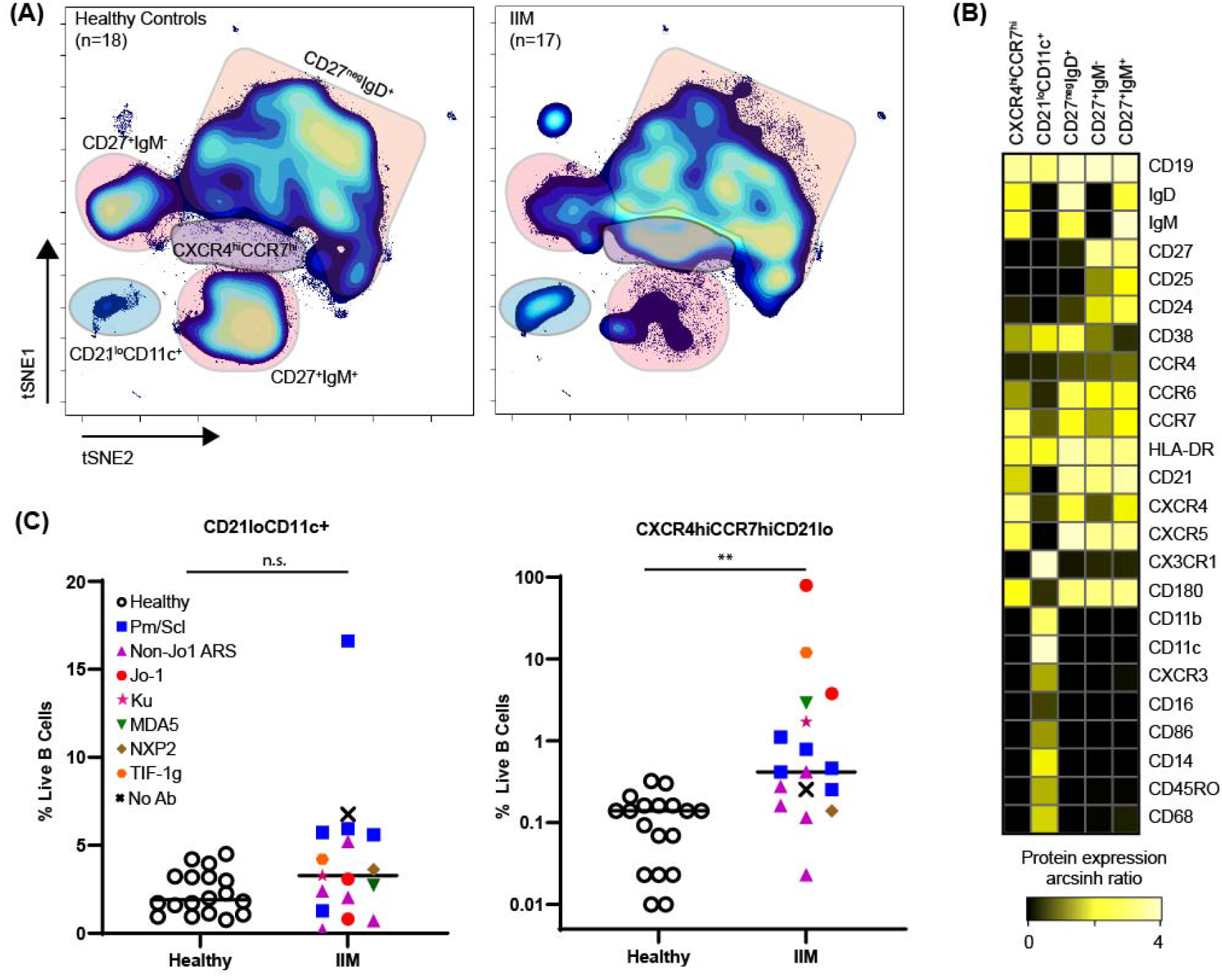
CD21loCD11c+ and CXCR4hiCCR7hi B cell subsets are increased amongst IIM patients. CD3-CD19+ B cells were mapped using tSNE. (A) Concatenated tSNE maps for all healthy controls (n=18) and IIM patients (n=17) showing traditional B cell subsets and two IIM-associated B cell subsets. (B) Expression heatmaps of representative markers for CD21loCD11c+ and CXCR4hiCCR7hi subsets displaying arcsinh ratio by table’s minimum of channel median. (C) Frequencies of CD21loCD11c+ and CXCR4hiCCR7hi subsets are shown for each donor where various myositis associated and specific autoantibodies are coded as follows: Statistical comparisons were performed using Mann-Whitney U tests, *p<0.05, **p<0.01, ***p<0.001, ***p<0.0001.

### High frequencies of abnormal CD19+ populations predict the presence of altered circulating CD4+ populations

As specific B cell subsets can affect T cell phenotype [13-15], we next investigated correlations between CD19+CXCR4hiCCR7hi and CD19+CD21loCD11c+ cell subsets and T cells. CITRUS is a fully automated machine learning algorithm that identifies cell populations statistically correlated with a particular biologic or clinical feature [24]. We created three mutually exclusive groups of IIM patients with high expression of CD19+CXCR4hiCCR7hi, CD19+CD21loCD11c+, or neither and compared to healthy controls (Supplemental Table S6). As shown in Figure 5, CITRUS identified increased abundance of CD4+CXCR4hiCD38hi naïve cells (Cluster A) in IIM patients defined by increased CD19+CXCR4hiCCR7hi cell frequency. CD4+CD27-PD1+ effector memory cells (Cluster B) were increased in the group with a high frequency of CD19+CD21loCD11c+ cells. CITRUS also identified several populations of T cells that were decreased in IIM patients (Supplemental Figure 2).

**Figure 5.**
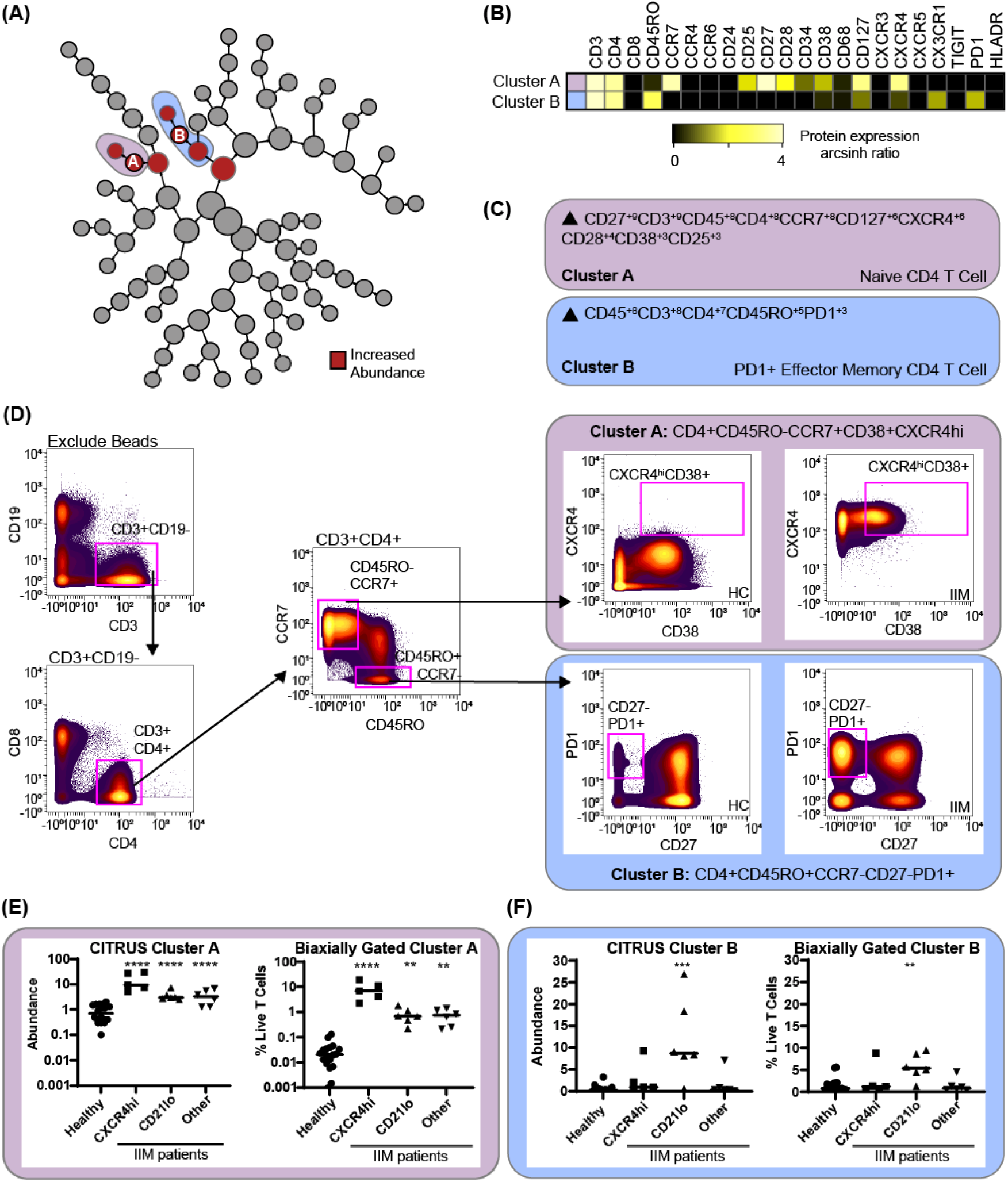
CITRUS identifies T cell clusters that are associated with abnormal CD19+CXCR4hiCCR7hi and CD19+CD21loCD11c+CD27-B cell populations. CD3+ T cells were clustered via CITRUS using four groups (healthy control, n=18, CXCR4hi, n=5, CD21lo, n=6, and other B cell phenotype, n=6) and the nearest shrunken centroid algorithm. (A) Increased abundance of clusters A and B are predicted by the presence of the CXCR4hi and CD21lo B cell subsets, respectively. (B) Expression heatmap of markers for Clusters A and B displaying arcsinh ratio by table’s minimum. (C) Algorithmic determination of biaxial gating scheme using marker enrichment modeling (MEM) for Clusters A and B. Scale 1-10. (D) Biaxial gating scheme derived based on MEM definition; Cluster A: CD4+CD45RO-CCR7+CXCXCR4hiCD38+, and Cluster B: CD4+CR7-CD45RO+CD27-PD1+. Representative healthy control (HC) and IIM patient plots are shown. (E) CITRUS determined abundance and corresponding biaxial gating frequency for Cluster A and (F) Cluster B are plotted for individual donors. If Kruskal-Wallis p<0.05, then post-hoc Mann-Whitney U tests were used to compare to healthy controls. *p<0.05, **p<0.01, ***p<0.001, ****p<0.0001.

To confirm this finding, marker enrichment modeling (MEM) computationally identified differentially expressed markers “enriched” in each cluster [27] to guide biaxial gating (Figure 5C-D). Biaxial population frequencies approximated the CITRUS-determined abundances (Figure 5E-F, Supplemental Table S7). CD19+CXCR4hiCCR7hi and CD19+CD21loCD11c+ population frequencies were plotted against the biaxially gated frequency of the CD4+CXCR4hiCD38+ and CD4+CD27-PD1+ subsets, respectively (Supplemental Figure 3A-B). We also calculated Spearman’s correlation coefficients (Supplemental Table S8) and confirmed a statistical correlation between CD19+CXCR4hiCCR7hi and CD4+CXCR4hiCD38+populations (r=0.62, p=0.009) and between CD19+CD21loCD11c+ CD4+CD27-PD1+ populations (r=0.60, p=0.01).

### An increased frequency of CD19+CXCR4hiCCR7hi cells predicts the increase of two myeloid populations

Using the previous IIM group assignments (Table S6), CITRUS clustered CD3-CD19-cells to correlate myeloid cell abundance with CD19+ cell phenotypes. Two clusters of increased abundance were predicted by having an increased frequency of CD19+CXCR4hiCCR7hi cells (Figure 6). MEM analysis identified Cluster C as a CD180+CXCR4+ classical monocyte and cluster C as a CXCR4-CD180-monocyte (Fig. 6C). The MEM-derived biaxial gating scheme did not have clear visual cut-offs and final biaxial gating (Figure 6D) required the use of a training data set. Thus, myeloid cluster increases were further validated by tSNE analysis (Supplemental Figure 4). Both the biaxial and tSNE frequencies approximated the CITRUS abundance (Figure 6E-F, Supplemental Figure 4D). Spearman’s correlation coefficients confirmed a statistical correlation of the CD19+CXCR4hiCCR7hi subset with both the biaxially gated classical monocyte (r=0.61, p=0.01) and the CXCR4-CD180-myeloid population (r=0.66, p=0.01).

**Figure 6.**
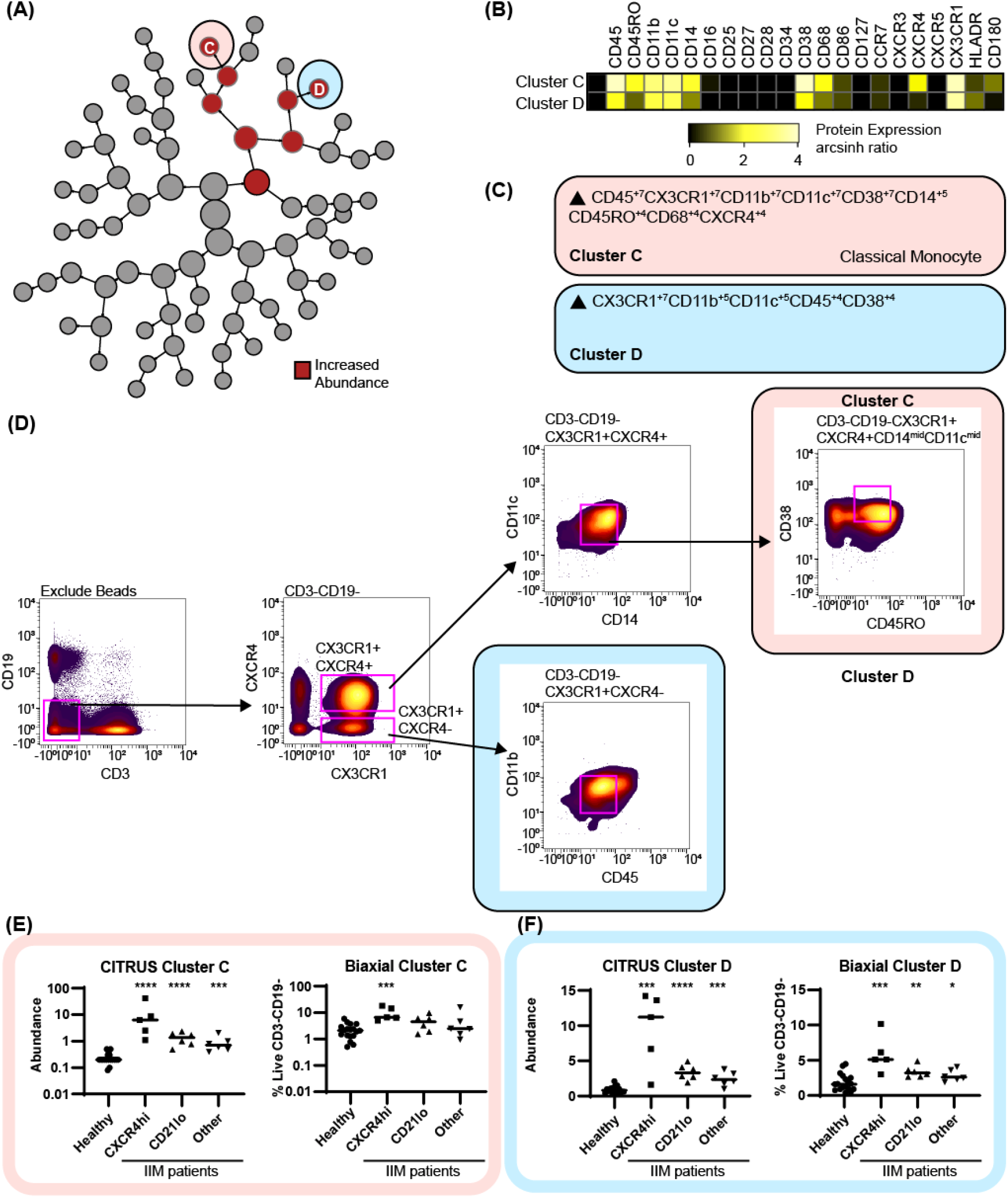
CITRUS identifies myeloid cell clusters that are increased in IIM. CD3-CD19-cells were clustered via CITRUS using four groups (healthy control, n=18, CXCR4hi, n=5, CD21lo, n=6, and other B cell phenotype, n=6) and the nearest shrunken centroid algorithm. (A) Increased abundance of Clusters C and D are increased across all IIM subsets. No clusters of decreased abundance were identified. (B) Expression heatmap of markers for Clusters C and D displaying arcsinh ratio by table’s minimum. (C) Algorithmic determination of biaxial gating scheme using marker enrichment modeling (MEM) for Clusters C and D. (D) Biaxial gating scheme derived based on MEM enrichment shown for a representative IIM patient. Additional tSNE gating is shown in Supplemental Figure 4. (E) CITRUS determined abundance and corresponding biaxial gating frequency for Cluster C and (F) Cluster D. If Kruskal-Wallis p<0.05, then post-hoc Mann-Whitney U tests were used to compare to healthy controls. *p<0.05, **p<0.01, ***p<0.001, ****p<0.0001.

## Discussion

Our key findings are: (1) significant heterogeneity exists in circulating PBMC phenotypes amongst IIM patients, (2) an abnormal CD19+CXCR4hiCCR7hi population correlates with CD4+CXCR4hiCD38+ T cell and CD14+CD16-HLADR-CXCR4hi classical monocyte populations, (3) an abnormal CD19+CD21loCD11c+ population correlates with a CD4+CD27-PD1+ T cell population, and (4) shared immunologic features amongst patients exist, including decreased memory B cells, low RP105/CD180 surface B cell expression, and depletion of CD3+CXCR3+ cells.

Several of our findings corroborate prior reports. Jo-1 patients were previously reported to have decreased memory B cells, as we also show [19]. While the increase of CD24hiCD38hi B cells did not reach statistical significance, some patients did have an increased frequency of this population as previously observed in juvenile dermatomyositis [20].

The different immune signatures observed amongst IIM patients in this report may have clinical ramifications. We identified a CD19+ CXCR4hiCCR7hi, CD3+CD4+CXCR4hiCD38hi, and CD14+CXCR4+ signature in a small number of IIM patients. Increased surface CXCR4 expression on circulating B and T cells has been associated with increased disease severity in SLE [35], and CXCR4+ infiltrates were previously identified in muscle biopsies in IIM patients [36]. Given these previous findings it is possible this signature is associated with disease severity, particularly considering the observation that 4/5 patients with the highest frequency of CD19+CXCR4hiCCR7hi cells were enrolled in the inpatient setting. CXCR4 can be upregulated by several stimuli [37-39], and the cause of CXCR4 upregulation in IIM is currently unclear.

The second CD19+ CD21loCD11c+ and CD4+CD27-PD1+ cell immune signature may represent a pro-fibrotic phenotype. These CD19+ cells may represent DN2 B cells previously reported in SLE [40]. In those studies, DN2 B cells were characterized as CD19+CD21loCD27-IgD-CXCR5-CD24-CD11c+Tbet+ and increased in African American Sm/RNP positive SLE patients with active disease. Transcriptionally, these cells appeared to be plasma cell precursors, but were also exquisitely sensitive to TLR7 stimulation. The association of DN2 cells with RNP autoantibody positivity is of particular interest as this, like Pm/Scl, is a systemic sclerosis overlap autoantibody. In our study, most patients with increased CD19+CD21loCD11c+ cells were Pm/Scl+. Alternatively, the numerous myeloid markers observed here raise the possibility that this CD19+ subset may not be of the B cell lineage [41, 42] or that the B cells may have acquired surface markers from follicular dendritic cells [43]. CD4+PD1+ T cells have been reported in a number of fibrotic diseases including subglottic stenosis [44], IPF, and sarcoidosis [45]. A trend towards increased frequency of CD4+PD1+ T cells has also been reported in systemic sclerosis [46].

Despite immunophenotypic differences, we identified some shared immunologic derangements across patients with IIM. First, decreased surface expression of RP105/CD180 could represent activation of the CD180 signaling cascade. CD180 is an alternative B cell signaling cascade capable of activating B cells either independently or synergistically with TLR9 or BCR engagement. CD180 is internalized after binding an unknown ligand, after which signaling occurs through an incompletely elucidated pathway [47]. Thus, we postulate decreased surface expression of CD180 may reflect increased CD180 internalization and signaling in IIM patients. We identified a general decrease in circulating T cells expressing CXCR3. This may be due to CXCR3+ T cell migration into sites of inflammation. This concept is supported by the fact that CXCR3+ T cells have been identified in the muscle biopsies of polymyositis, dermatomyositis, and inclusion body myositis patients [48]. Interferon gamma and tumor necrosis factor alpha, which are increased in IIM, can induce secretion of the CXCR3 ligand CXCL10 by muscle fibers [49]. Of interest, CXCR3 ligands, CXCL9 and CXCL10, are increased in the serum of Jo1+ and SRP+ myositis patients [50] and CXCL10 is a validated disease activity biomarker for juvenile dermatomyositis [51]. While many patients in this cohort were on corticosteroids, CXCR3+ T cells were unchanged in asthmatics treated with moderate prednisolone for two weeks [52], making it less likely that reduced circulating CXCR3+ T cells is due solely to steroid use.

This study has several strengths. First, CyTOF facilitated simultaneous evaluation of multiple PBMC subsets using minimal sample with single cell resolution. Second, incorporating unsupervised analysis with traditional biaxial gating confirmed prior findings while also identifying novel subsets in a comprehensive and minimally biased way. Third, hospitalized patient inclusion permitted us to investigate the entire spectrum of disease severity from mildly symptomatic outpatients to critically ill individuals requiring extracorporeal membrane oxygenation. We acknowledge this is a single center cohort with substantial serologic heterogeneity and an overrepresentation of ILD and validation in a larger cohort will be required.

In conclusion, we show that significant immunophenotypic heterogeneity exists within IIM and identify two distinct immune signatures within this cohort. We find shared characteristics of the IIM immunophenotype across the entire IIM spectrum, demonstrating that B cell surface RP105/CD180 expression and circulating CXCR3+ T cells are decreased. Future work is needed to determine the clinical significance of these novel immune signatures.

## Data Availability

All data is included in either the main manuscript or supplemental materials.

## Acknowledgements

This work was supported by CTSA award No. UL1TR000445 (EMW, LJC) from the National Center for Advancing Translational Sciences, the National Institutes of Health T32HL087738 (EMW), KL2TR002245 (EMW), K12HD043483 (RHB), T32AR0590139 (JJY), K00-CA212447 (TB), R01 CA226833 (JMI), U54 CA217450 (JMI), The Myositis Association (EMW), Myositis UK (EMW), Vanderbilt Faculty Research Scholars Award (EMW), Vanderbilt Human Immunology Discovery Initiative, and the Porter Family Fund for Autoimmunity Research. The contents are solely the responsibility of the authors and do not necessarily represent official views of the National Center for Advancing Translational Sciences or the National Institutes of Health.

## Competing Interests

EMW receives research funding from Boehringer-Ingelheim and is a member of their myositis ILD advisory board. JMI was a co-founder and a board member of Cytobank Inc. and has engaged in sponsored research with Incyte Corp, Janssen, Pharmacyclics. No other author reports a competing interest.

## Key Messages

What is already known about this subject?

- Prior studies evaluated individual phenotypic differences in B, T, and monocyte populations.

What does this study add?

- Simultaneous, comprehensive immunophenotyping of adaptive and innate PBMC subsets identified two distinct immune signatures comprised of a B and T cell subset pair.

- All peripheral blood CXCR3^+^ T cell subsets were depleted in IIM patients.

How might this impact on clinical practice or future developments?

- Immune signatures may correlate with clinical manifestations and suggest novel targeted therapies.

## Abbreviations

(ARS): anti-tRNA synthetase syndrome
(BCR): B cell receptor
(CITRUS): cluster identification, characterization, and regression
ILD): interstitial lung disease
(CyTOF): mass cytometry by time of flight
(IIM): idiopathic inflammatory myopathies
(MEM): marker enrichment modeling
(MHCII): major histocompatibility complex class II
(MFI): Median Mass Intensities
(MMI): mean fluorescence intensity
(PBMCs): peripheral blood mononuclear cells
(tSNE): t-distributed stochastic neighbor embedding

## Notes

### Author Declarations

VUMC IRB 141415

