## Supplemental Materials for "High-dimensional analysis reveals abnormal B cell subsets associated with specific changes to circulating T and myeloid cell populations in patients with idiopathic inflammatory myopathies"

### **Supplemental Methods**

#### **Cytometry by time of flight (CyTOF) staining and data acquisition**

Individual PBMC cryotubes were thawed in 10 mL warm PBS w/o calcium or magnesium (Gibco, Life Technologies, Grand Island, NY), pelleted by centrifugation and washed twice with 10 mL PBS. Cells were transferred to a 96-well plate for staining. Cells were incubated first with a viability reagent (200nM cisplatin-198, Fluidigm) in phosphate buffered saline (PBS) for 5 minutes; the reaction was quenched using 125 µL PBS/1% bovine serum albumin (BSA). Clones used for staining are shown in table S1. Cells were washed twice with PBS/1% BSA. Cells were resuspended in 100 µL surface stain master mix and 30 µL primary surface master mix (Table S1) and incubated for 30 minutes at room temperature (RT). 50 µL of PBS/1%BSA was added to each well, the cells were pelleted by centrifugation, and washed once with PBS/1% BSA. The cells were resuspended in 100 µL secondary surface master mix for 30 minutes. Cells were pelleted by centrifugation and washed once with PBS without BSA. Cells were then fixed with 100 µL 1.6% paraformaldehyde, incubated for 20 minutes and washed with PBS. Cells were permeabilized using Ebioscience FoxP3 fix/perm buffer (Thermofisher) for 45 minutes at RT. Cells were pelleted by centrifugation and washed in fix/perm buffer prior to resuspension in 100 µL primary intracellular master mix (Table S2) for 30 minutes at RT. Cells were pelleted by centrifugation and washed with fix/perm buffer before resuspending in secondary intracellular mastermix for 30 minutes at RT. Cells were pelleted by centrifugation, washed with fix/perm buffer, and resuspended in 1.6% paraformaldehyde with 6.25 nM intercalator overnight at 4 degrees. Data was collected within 72h of staining.

On the day of data acquisition, cells were washed twice with PBS without calcium or magnesium and once with 2 mL milliQ water. Cell concentration was adjusted to ~500,000 cells/mL with milliQ water and 10% volume of equilibration beads (Fluidigm Sciences, Sunnyvale, CA) was added to the cell suspension. Cells were filtered immediately before injection into the mass cytometer using a 35 µm nylon mesh cell-strainer cap (BD Biosciences). Data was acquired using a CyTOF Helios (Fluidigm Sciences, Sunnyvale, CA) and CyTOF software (version 6.7.1014) at the Vanderbilt University Medical Center Mass Cytometry Center of Excellence. Dual count calibration and noise reduction were applied during the acquisition; 100,000-400,000 events were collected per sample.

| **Table S1.** Direct surface staining antibody clones | | |
| --- | --- | --- |
| **Metal** | **Target** | **Clone and Source** |
| 89Y | CD45 | HI30 (Fluidigm) |
| 141Pr | CCR6 | G034E3 (Fluidigm |
| 142Nd | CD19 | HIB19 (Fluidigm) |
| 144Nd | CD11b | ICRF44 (Fluidigm) |
| 145Nd | CD4 | RPA-T4 (Fluidigm) |
| 146Nd | IgD | IA6-2 (Fluidigm) |
| 147Sm | CD11c | Bu15 (Fluidigm) |
| 148Nd | CD16 | 3G8 (Fluidigm) |
| 149Sm | CD127 | A019D5 (Fluidigm) |
| 150Nd | CD86 | IT2.2 (Fluidigm) |
| 151Eu | HLA-DR | G46-6 (Fluidigm) |
| 152Sm | CD21 | BL13 (Fluidigm) |
| 153Eu | CXCR5 | RF8B2 (Fluidigm) |
| 154Sm | TIGIT | MBSA43 (Fluidigm) |
| 155Gd | PD-1 | EH12.2H7 (Fluidigm) |
| 156Gd | CXCR3 | G025H7 (Fluidigm) |
| 158Gd | CCR4 | L291H4 (Fluidigm) |
| 159Tb | CCR7 | G043H7 (Fluidigm) |
| 160Gd | CD14 | M5E2 (Fluidigm) |
| 162Dy | CD27 | L128 (Fluidigm) |
| 163Dy | CD34 | 581 (Fluidigm) |
| 164Dy | CD45RO | UCHL1 (Fluidigm) |
| 166Er | CD24 | ML5 (Fluidigm) |
| 167Er | CD38 | HIT2 (Fluidigm) |
| 168Er | CD8 | SKI1 (Fluidigm) |
| 169Tm | CD25 | 2A3 (Fluidigm) |
| 170Er | CD3 | UCHT1 (Fluidigm) |
| 172Yb | IgM | MHM-88 (Fluidigm) |
| 173Yb | CXCR4 | 12G5 (Fluidigm) |

| **Table S2**. Primary and Secondary Antibodies | | | |
| --- | --- | --- | --- |
| **PRIMARY SURFACE MASTERMIX** | | | |
| **Metal** | **Target** | **Clone and Source** | |
| -- | CX3CR1-biotin | 2A91 (Biolegend) | |
| -- | CD28- APC | CD28.2 (Biolgend) | |
| **SECONDARY SURFACE MASTERMIX** | | | |
| **Metal** | **Target** | | **Clone and Source** |
| 143Nd | Anti-biotin | | ID4-C5 (Fluidigm) |
| 176Yb | Anti-APC | | APC003 (Fluidigm) |
| **PRIMARY INTRACELLULAR MASTERMIX** | | | |
| **Metal** | **Target** | | **Clone and Source** |
| 171Yb | CD68 | | Y1/82A (Fluidigm) |
| -- | Rabbit anti-BTK | | D3H5 (CellSignaling) |
| **SECONDARY INTRACELLULAR MASTERMIX** | | | |
| **Metal** | **Target** | | **Clone and Source** |
| 174Yb | anti-FITC | | FIT-22 (Fluidigm) |
| 175Lu | Goat anti-rabbit | | Polyclonal (Fluidigm) |

#### **Flow Cytometry Staining and Data Acquisition**

Cryopreserved PBMCs of 6 healthy controls and 6 SSc-ILD were thawed in 10 mL of PBS w/o calcium or magnesium, pelleted by centrifugation, resuspended in fluorescence-activated cell sorting (FACS) buffer containing fetal bovine serum and sodium azide, and washed once more with FACS buffer. Cells were transferred to a 96-well plate for staining. After incubation with 5 μL Fc Block (BD Biosciences, San Jose, CA) in 45μL FACS buffer for 10 minutes on ice, surface mastermix (Table S4) was added for 30 minutes on ice without centrifugation prior to adding Live/Dead 700 for an additional 5 minutes. Cells were pelleted by centrifugation, washed twice with FACS buffer, and resuspended in eBioscience FoxP3 Fix/Perm solution (ThermoFisher Scientific, USA) for 30 minutes, pelleted by centrifugation, and washed with FoxP3 Fix/Perm buffer. After pelleting by centrifugation, cells were re-suspended in intracellular mastermix (Table S3) in FoxP3 Fix/Perm buffer for 30 min at RT, pelleted by centrifugation and washed twice with FoxP3 Fix/Perm buffer prior to resuspension in 1% PFA with 2 mM EDTA and transferring FACS tubes for data acquisition. All data was acquired on a BD LSRII Fortessa instrument.

| **Table S3.** Extracellular and Intracellular Flow Antibodies | | | |
| --- | --- | --- | --- |
| **EXTRACELLULAR MASTERMIX** | | | |
| **Fluor** | **Target** | **Clone and Source** | |
| APC | CD180 | MHR73-11 (Biolegend) | |
| BV785 | CD27 | O323 (Biolegend) | |
| BV510 | CD3 | OKT3 (Biolegend) | |
| BV510 | CD14 | M5E2 (Biolegend) | |
| BV510 | CD16 | 3G8 (Biolegend) | |
| BUV395 | CD19 | SJ25C1 (BD) | |
| **INTRACELLULAR MASTERMIX** | | | |
| **Fluor** | **Target** | | **Clone and Source** |
| PE | CD180 | | MHR73-11 (Biolegend) |

#### **CyTOF data analysis:**

viSNE/tSNE Analysis

All tSNE and CITRUS analyses were conducted through the Cytobank software suite. Advanced analyses with viSNE were utilized to generate tSNE plots. Detailed settings for all tSNE/viSNE analyses are shown in Table S4.

| **Table S4.** Settings for tSNE/viSNE analyses | | | | | | |
| --- | --- | --- | --- | --- | --- | --- |
|  | Figure 1 (PBMC viSNE) | | | Figure 4 (CD19+ viSNE) | | |
| Gated population | Exclude beads | | | CD19+ Cells | | |
| Total #Events | 750,003 | | | 155,556 | | |
| Sampling | Proportional  (minimum 5348 events/sample) | | | Equal  4321 events/sample | | |
| Perplexity | 90 | | | 75 | | |
| Theta | 0.5 | | | 0.5 | | |
| #Iterations | 7500 | | | 7500 | | |
|  | CCR6 | CD19 | CXC3CR1 | CCR6 | CD19 | CX3CR1 |
|  | CD11b | CD4 | IgD | CD11b | CD4 | IgD |
|  | CD11c | CD16 | CD127 | CD11c | CD16 | CD127 |
|  | CD86 | HLA-DR | CD21 | CD86 | HLA-DR | CD21 |
|  | CXCR5 | TIGIT | PD1 | CXCR5 | TIGIT | PD1 |
|  | CXCR3 | CCR4 | CCR7 | CXCR3 | CCR4 | CCR7 |
|  | CD14 | CD27 | CD34 | CD14 | CD27 | CD34 |
|  | CD45RO | CD180 | CD24 | CD45RO | CD180 | CD24 |
|  | CD38 | CD8 | CD25 | CD38 | CD25 | CD68 |
|  | CD3 | CD68 | IgM | IgM | CXCR4 | CD28 |
|  | CXCR4 | CD28 | CD45 |  |  |  |

CITRUS Analysis:

CITRUS analysis was performed within the Cytobank software suite. B cell immunophenotypes were assigned based on the viSNE analysis shown in Figure 4. The five patients with the highest frequency of CXCR4hiCCR7hi B cells were assigned to the CXCR4hi group. The six patients with the highest frequency of CD19+CD21loCD11c+ cells were assigned to the CD21lo group, and the remaining patients were assigned as “Other.” Exact settings of the CITRUS algorithm are shown in Table S5.

| **Table S5.** Settings for CITRUS | | | | | | |
| --- | --- | --- | --- | --- | --- | --- |
|  | Figure 5 (T cell CITRUS) | | | Figure 6 (Myeloid cell CITRUS) | | |
| **Gated population** | CD3+ Cells | | | CD3-CD19- Cells | | |
| **Association Model** | Nearest Shrunken Centroid (PAMR) | | | Nearest Shrunken Centroid (PAMR) | | |
| **Total #Events** | 262,500 | | | 262,500 | | |
| **Events per sample** | 7,500 | | | 7,500 | | |
| **Minimum cluster size** | 2% (5250 events) | | | 2% (5250) | | |
| **Cross Validation Folds** | 5 | | | 5 | | |
| **False discovery rate** | 1% | | | 1% | | |
| **Markers** | CX3CR1 | CD11b | CD4 | CCR6 | CX3CR1 | CD11b |
|  | CD11c | CD16 | CD127 | CD4 | CD11c | CD16 |
|  | CD86 | HLA-DR | CD21 | CD127 | CD86 | HLA-DR |
|  | CD21 | CXCR5 | TIGIT | CD21 | CXCR5 | TIGIT |
|  | PD-1 | CXCR3 | CCR4 | PD-1 | CXCR3 | CCR4 |
|  | CCR7 | CD14 | CD27 | CCR7 | CD14 | CD27 |
|  | CD34 | CD45RO | CD24 | CD38 | CD8 | CD25 |
|  | CD38 | CD8 | CD25 | CD68 | IgM | CXCR4 |
|  | CD68 | CXCR4 | CD28 | CD28 |  |  |

**Marker Enrichment Modeling (MEM) Analysis**

**Determination of Biaxial gating scheme:**

For Clusters A and B, biaxial gating was performed using easily visualized differences in contour maps using MEM markers. For Clusters C and D, clear breakpoints were not apparent. FCS files corresponding to CITRUS clusters C and D were exported, concatenated, and loaded back into Cytobank. Using the gating editor, the concatenated files were used to train the biaxial gating scheme and then these gates were applied to the original (pre-CITRUS) FCS files.

### **Supplemental Figures**

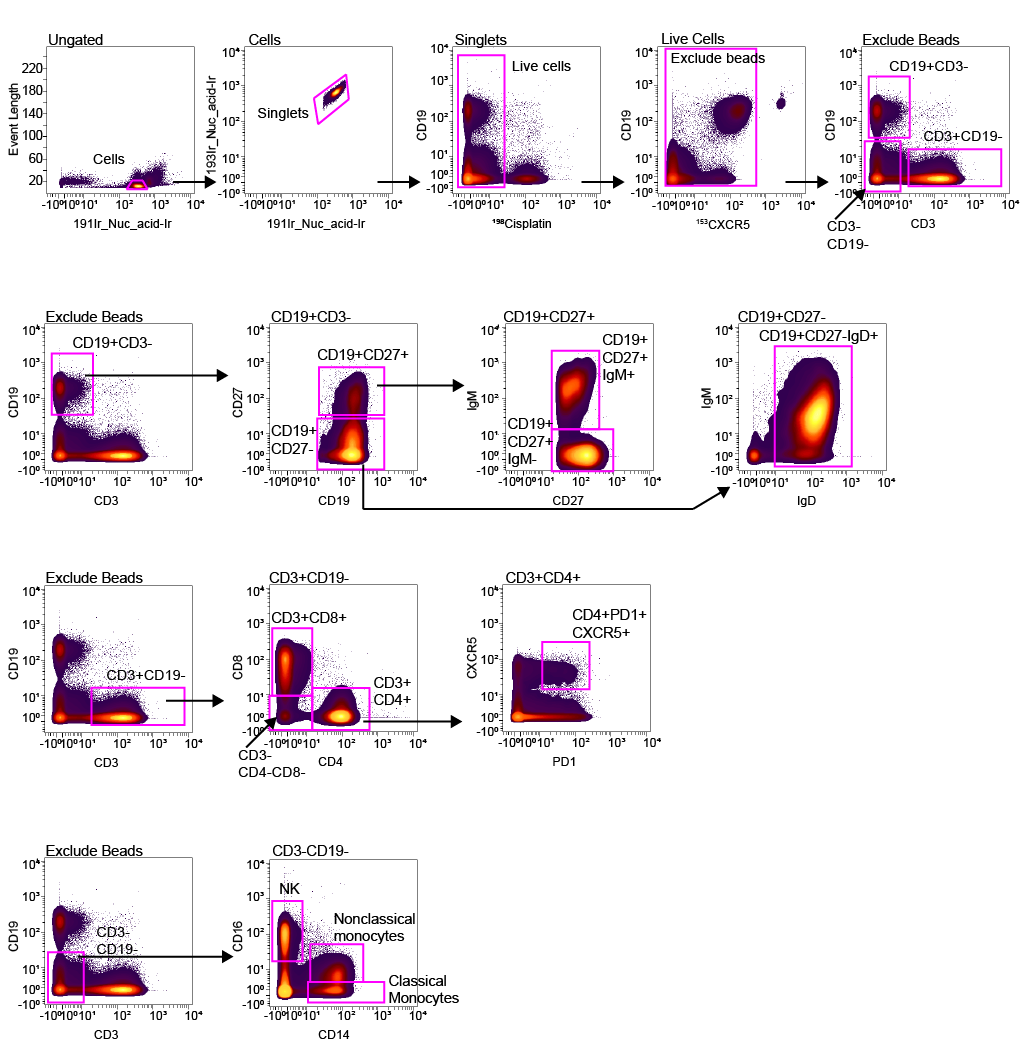

**Supplemental Figure S1**. Biaxial gating scheme for major PBMC subsets. Biaxial gating schemes were designed based on the Human Immunology Project. (PMCID 3409649)

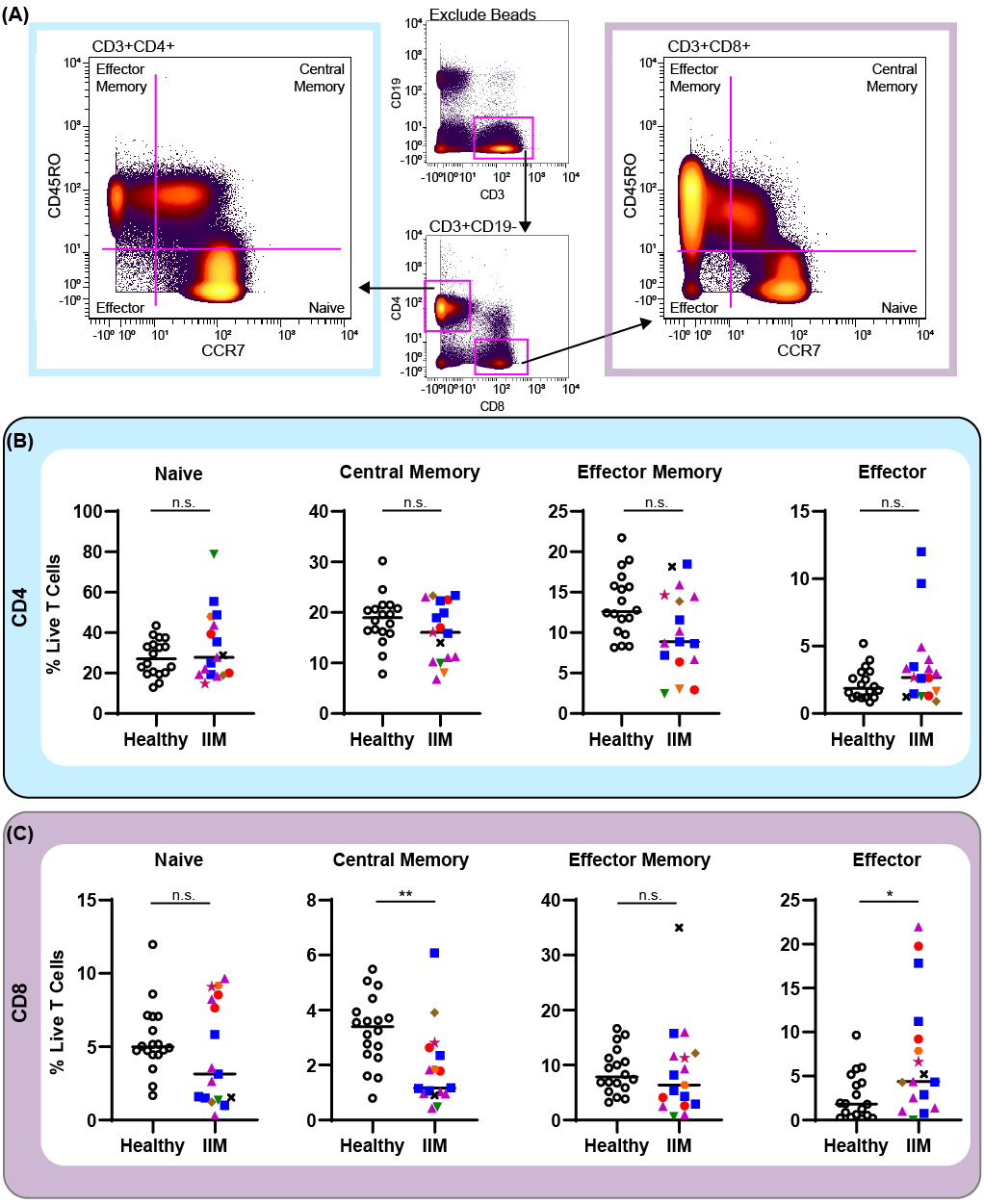

**Supplemental Figure 2.** Biaxial gating of CD4+ and CD8+ T cells to quantify naive, central memory, effector memory, and effector populations. Biaxial gating was performed on 18 healthy controls and 17 IIM patients. (A) Biaxial gating scheme to identify naive, central memory, effector memory, and effector populations. (B) Quantification of CD4+ and (C) CD8+ naive, central memory, effector memory, and effector populations. Mann-Whitney U tests were used for statistical comparison. *p<0.05, **p<0.01, ***p<0.001, ****p<0.0001

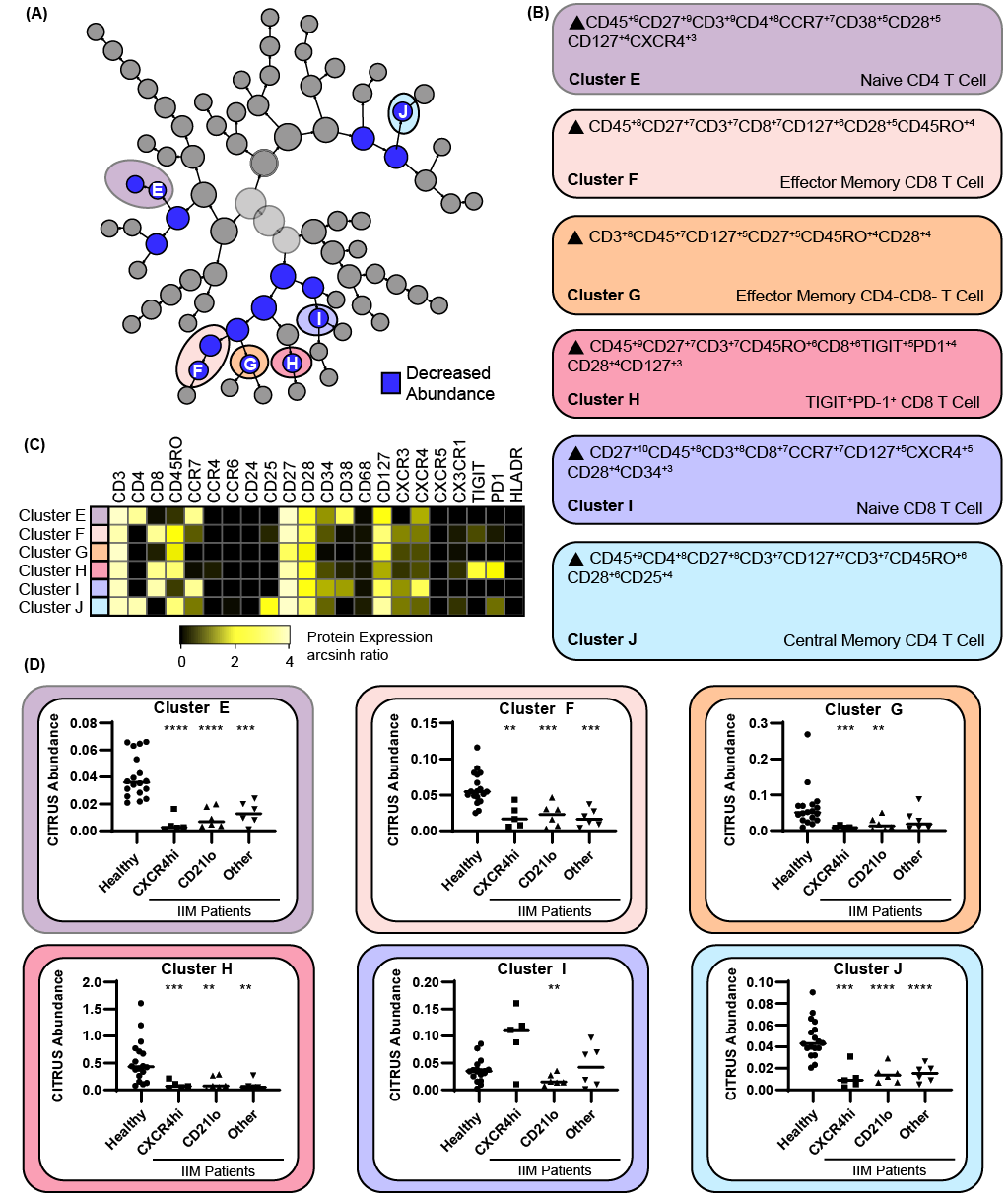

**Supplemental Figure 3.** CITRUS identifies T cell clusters that are decreased in IIM compared to heathy controls. CD3+ T cells were clustered via CITRUS using four groups (healthy control, n=18, CXCR4hi, n=5, CD21lo, n=6, and other B cell phenotype, n=6) with using the nearest shrunken centroid algorithm. (A) Six T cell clusters with decreased abundance were identified by CITRUS. (B) Algorithmic determination of surface marker expression of clusters A-F using marker enrich-ment modeling (MEM). (C) Expression heatmap for clusters A-F displaying arcsinh ratio by table’s minimum of the channels median. (D) CITRUS determined abundance for clusters A through E. If Kruskal-Wallis p<0.05, then post-hoc Mann-Whitney U tests were used to compare to healthy controls. *p<0.05, **p<0.01, ***p<0.001, ****p<0.0001.

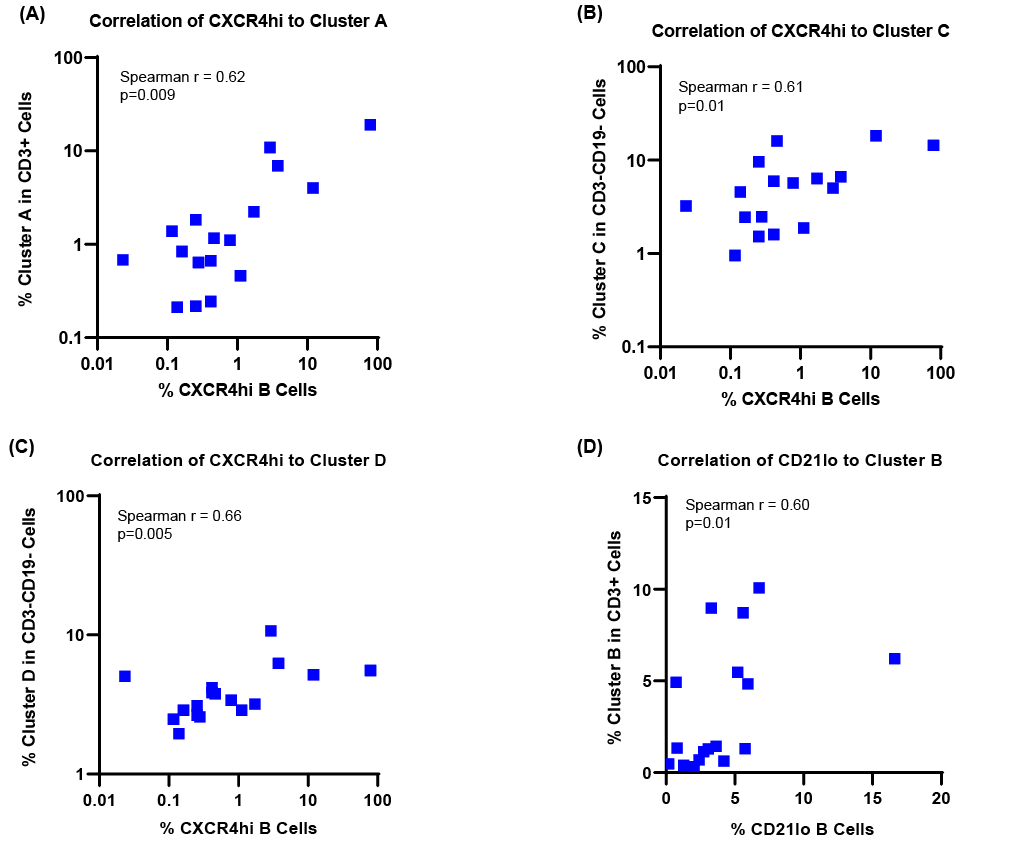

**Supplemental Figure 4.** B cell subsets statistically correlate with T and myeloid subsets identified through CITRUS. Frequency of CXCR4hi B cells identified in the Figure 4 tSNE analysis plotted against the biaxial populations corresponding to CITRUS (A) cluster A, (B) cluster C, and (C) cluster D shown in Figures 5-6 with corresponding Spearman’s correlation coefficient. (D) Frequency of biaxial population corresponding to CITRUS cluster B against the frequency of CD19+CD21loCD11c+ cells with corresponding Spearman’s correlation coefficient.

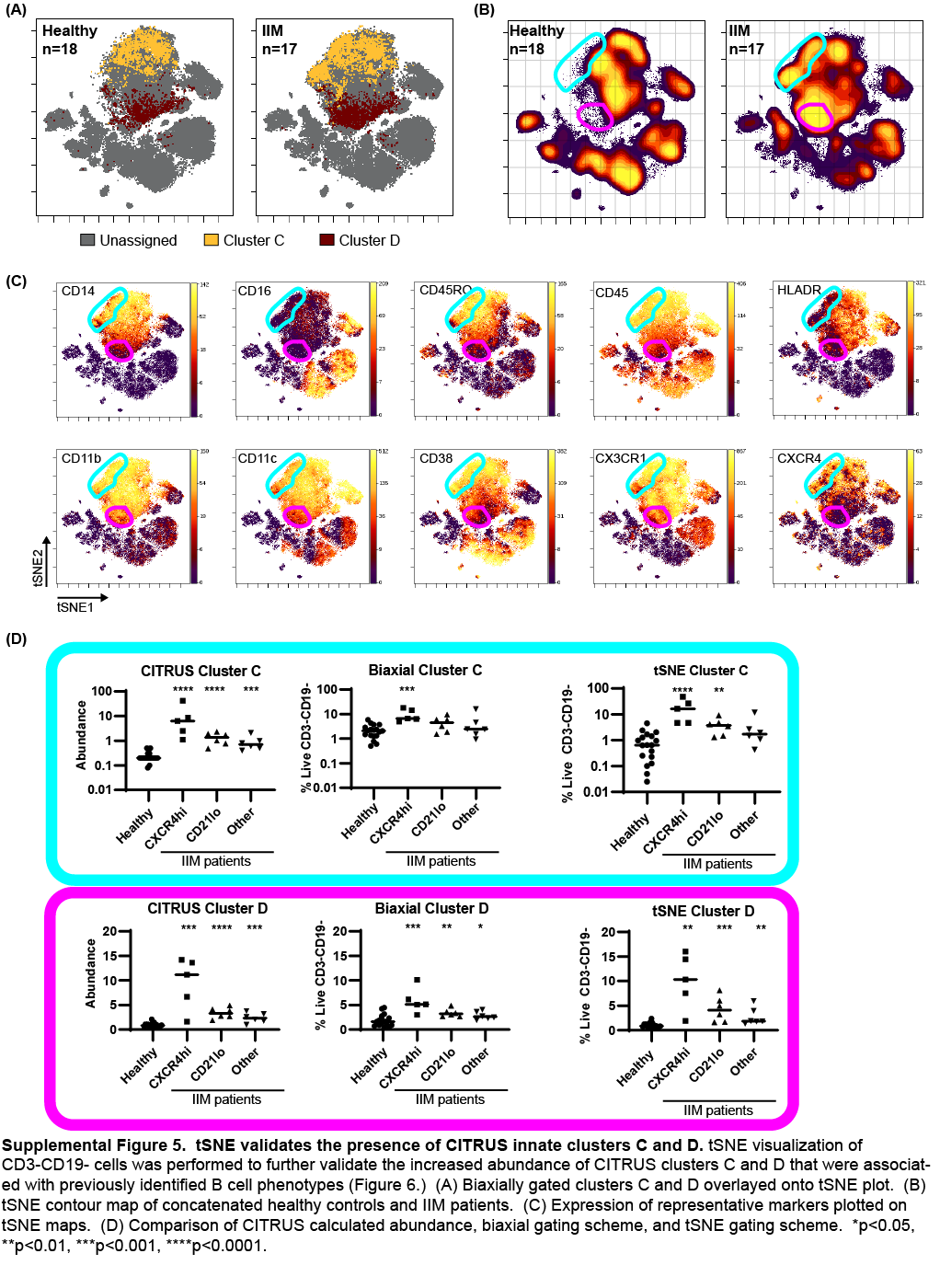

**Supplemental Figure 5.** tSNE validates the presence of CITRUS innate clusters C and D. tSNE visualization of CD3-CD19- cells was performed to further validate the increased abundance of CITRUS clusters C and D that were associat-ed with previously identified B cell phenotypes (Figure 6.) (A) Biaxially gated clusters C and D overlayed onto tSNE plot. (B) tSNE contour map of concatenated healthy controls and IIM patients. (C) Expression of representative markers plotted on tSNE maps. (D) Comparison of CITRUS calculated abundance, biaxial gating scheme, and tSNE gating scheme. *p<0.05, **p<0.01, ***p<0.001, ****p<0.0001.

### **Supplemental Data Tables**

| **Supplemental Table S6.** Detailed clinical characteristics of IIM patients | | | | | | | | |  |
| --- | --- | --- | --- | --- | --- | --- | --- | --- | --- |
| **Sample ID** | **Age** | **Gender** | **Race** | **Estimated disease duration** | **Antibody** | **Disease manifestations** | **Medications at time of blood draw** | **Enrollment location** | **Meets IIM Critieria** |
| 1 | 61-66 | M | Caucasian | 0.7 | MDA5 | Mechanic’s hands, ILD | Methylprednisolone 1000 mg IV (day 3) | ICU | No |
| 2 | 69-74 | M | Caucasian | 0.8 | NXP2 | Proximal muscle weakness, ILD | None | Clinic | No |
| 3 | 35-40 | F | African American | 3.0 | PL12/Ro52 | ILD, dysphagia, Mechanic’s hands | None | Clinic | Yes |
| 4 | 70-74 | F | Caucasian | 0.9 | PL12/Ro52 | ILD, GERD | Prednisone 10mg | Clinic | No |
| 5 | 46-50 | M | Caucasian | 4.0 | Jo1 | Shawl sign, proximal muscle weakness, ILD, arthritis | Methylprednisolone 1000 mg IV (day 2) | ICU | Yes |
| 6 | 53-57 | M | Caucasian | 0.6 | Pm/Scl | Mechanic’s hands, ILD, arthritis | Methotrexate 15 mg/week | Clinic | No |
| 7 | 45-49 | M | African American | 1.0 | Jo1/Ro52 | Mechanic’s hands, ILD, proximal muscle weakness | None | Inpatient floor | Yes |
| 8 | 75-79 | F | Caucasian | 2.2 | Pm/Scl | Arthritis, proximal muscle weakness, ILD, pulmonary hypertension | Prednisone 40mg/day | Clinic | Yes |
| 9 | 59-63 | M | African American | 7.1 | PL7 | ILD, GERD | None | Clinic | No |
| 10 | 71-75 | F | Caucasian | 1.3 | Tif1g | Proximal muscle weakness, Gottron’s, heliotrope | Prednisone 60 mg/day | Inpatient floor | Yes |
| 11 | 49-54 | F | Caucasian | 0.9 | Ku/Ro52 | Gottron’s papules, ILD | Prednisone 70 mg/day | Clinic | Yes |
| 12 | 51-55 | M | Caucasian | 0.3 | Pm/Scl | ILD, proximal muscle weakness | Prednisone 60 mg/day | Clinic | Yes |
| 13 | 35-39 | F | Caucasian | 0.7 | EJ/Ro52 | ILD | None | Clinic | No |
| 14 | 59-64 | M | Caucasian | 0.8 | Pm/Scl | Proximal muscle weakness, dysphagia | Prednisone 60 mg/day | Inpatient Floor | Yes |
| 15 | 52-56 | F | Caucasian | 0.9 | Pm/Scl | Heliotrope, Gottron’s, mechanic’s hands, ILD, proximal muscle weakness | None | Clinic | Yes |
| 16 | 60-64 | M | Caucasian | 0.9 | PL7 | Heliotrope rash, Gottron’s, dysphagia, ILD | None | Clinic | Yes |
| 17 | 65-69 | F | African American | 0.52 | None | Proximal muscle weakness, + muscle bx | None | Inpatient Floor | Yes |

| **Supplemental Table S7.** CITRUS group assignments with CXCR4hiCCR7hi and CD21loCD11c+ frequencies | | | |
| --- | --- | --- | --- |
| Group Assignment | Sample ID | %CXCR4hiCCR7hi  in live CD19+ | %CD21loCD11c+  in live CD19+ |
| CXCR4hi | 1 | **2.916** | 2.0366 |
|  | 5 | **79.4261** | 0.81 |
|  | 7 | **3.7723** | 3.078 |
|  | 10 | **11.988** | 4.212 |
|  | 11 | **1.7126** | 3.2863 |
| CD21lo | 6 | 0.7869 | **5.6006** |
|  | 8 | 1.1109 | **5.9477** |
|  | 14 | 0.4166 | **16.6165** |
|  | 15 | 0.2546 | **5.7394** |
|  | 16 | 0.0231 | **5.2071** |
|  | 17 | 0.2546 | **6.7577** |
| Other | 2 | 0.1389 | 3.6334 |
|  | 3 | 0.2777 | 0.1851 |
|  | 4 | 0.162 | 2.4069 |
|  | 9 | 0.4166 | 2.7308 |
|  | 11 | 0.4629 | 1.2729 |
|  | 13 | 0.1157 | 0.7174 |
| Healthy Controls |  | 0.0694 | 1.134 |
|  |  | 0.0231 | 1.5969 |
|  |  | 0.3009 | 0.7637 |
|  |  | 0.162 | 1.7126 |
|  |  | 0.0926 | 1.0646 |
|  |  | 0 | 1.9903 |
|  |  | 0.1389 | 4.212 |
|  |  | 0.0231 | 0.9489 |
|  |  | 0.2083 | 3.1937 |
|  |  | 0 | 1.7126 |
|  |  | 0.162 | 4.5128 |
|  |  | 0.162 | 1.8051 |
|  |  | 0.324 | 3.24 |
|  |  | 0.1389 | 0.9489 |
|  |  | 0.0694 | 3.9574 |
|  |  | 0.1389 | 3.1937 |
|  |  | 0.1389 | 2.9854 |
|  |  | 0.0231 | 2.268 |

| **Supplemental Table S8.** Means, standard deviations, and p values for CITRUS clusters and biaxially gated populations | | | | | | | | |
| --- | --- | --- | --- | --- | --- | --- | --- | --- |
|  | Healthy Controls | IIM Patients by Predominant B Cell Phenotype | | | Statistical Comparisons | | | |
|  |  | CXCR4hi | CD21lo | Other | Kruskal-Wallis ANOVA | Healthy v. CXCR4hi | Healthy v. CD21lo | Healthy v. Other |
| Cluster A CITRUS abundance^†^ | 0.01 ± 0.01 | 0.16 ± 0.12 | 0.03 ± 0.01 | 0.04 ± 0.02 | <0.0001 | <0.0001 | 0.001 | 0.002 |
| Cluster A biaxial gating^‡^ | 0.03 ± 0.03% | 8.6 ± 6.7% | 0.8 ± 0.6 | 0.7 ± 0.5 | <0.0001 | <0.0001 | <0.0001 | <0.0001 |
| Cluster B CITRUS abundance^†^ | 0.01 ± 0.02 | 0.03 ± 0.04 | 0.12 ± 0.09 | 0.02 ± 0.03 | 0.01 | 0.36 | 0.0006 | 0.94 |
| Cluster B biaxial gating^‡^ | 1.5 ± 1.6% | 2.4 ± 3.6% | 5.7 ± 3.0% | 1.4 ± 1.6% | 0.03 | 0.97 | 0.003 | 0.62 |
| Cluster C CITRUS abundance^§^ | 0.002 ± 0.001 | 0.12 ± 0.17 | 0.01 ± 0.007 | 0.009 ±0.006 | <0.0001 | <0.0001 | <0.0001 | 0.0001 |
| Cluster C biaxial gating^⁑^ | 2.3 ± 1.4% | 10.1 ± 5.9% | 4.7 ± 3.1% | 4.7 ± 5.7% | 0.003 | 0.0001 | 0.07 | 0.27 |
| Cluster D CITRUS abundance^§^ | 0.01 ± 0.005 | 0.09 ± 0.05 | 0.03 ± 0.01 | 0.02 ± 0.01 | <0.0001 | 0.0001 | <0.0001 | 0.0007 |
| Cluster D biaxial gating^⁑^ | 1.9 ± 1.2% | 5.9 ± 2.6% | 2.5 ± 4.2% | 2.9 ± 0.8% | 0.0001 | 0.0004 | 0.007 | 0.04 |
| Cluster E CITRUS abundance^†^ | 0.04 ± 0.02 | 0.005± 0.006 | 0.009 ± 0.007 | 0.01 ± 0.008 | <0.0001 | <0.0001 | <0.0001 | 0.0001 |
| Cluster F CITRUS abundance^†^ | 0.06 ± 0.02 | 0.02 ± 0.02 | 0.02 ± 0.02 | 0.02 ± 0.01 | 0.0002 | 0.002 | 0.001 | 0.0001 |
| Cluster G CITRUS abundance^†^ | 0.06 ± 0.06 | 0.009 ± 0.005 | 0.02 ± 0.02 | 0.03 ± 0.03 | 0.002 | 0.0001 | 0.0074 | 0.06 |
| Cluster H CITRUS abundance^†^ | 0.06 ± 0.04 | 0.02 ± 0.01 | 0.02 ± 0.02 | 0.03 ± 0.02 | 0.0007 | 0.0004 | 0.004 | 0.007 |
| Cluster I CITRUS abundance^†^ | 0.05 ± 0.02 | 0.10 ± 0.06 | 0.02 ± 0.01 | 0.4 ± 0.4 | 0.02 | 0.06 | 0.001 | 0.78 |
| Cluster J CITRUS abundance^†^ | 0.05 ± 0.02 | 0.01 ± 0.02 | 0.02 ± 0.008 | 0.02 ± 0.008 | <0.0001 | 0.0002 | <0.0001 | <0.0001 |
| ^†^ abundance of all CD3+ cells (T) cells  ^‡^ frequency of all CD3+ cells (T) cells  ^§^ abundance of all CD3-CD19- (innate) cells  ^⁑^ frequency of all CD3-CD19- (innate) cells | | | | | | | | |

| **Supplemental Table S9.** Spearman’s correlation coefficients and p values for biaxially gated CITRUS clusters A-D | | | | |
| --- | --- | --- | --- | --- |
| B cell population | Cluster A | Cluster B | Cluster C | Cluster D |
| CD19+CXCR4hiCCR7+ | **0.6221**** | -0.2466 | **0.6061*** | **0.6581**** |
| CD19+CD21loCD11c+ | -0.3587 | **0.5980*** | 0.0098 | -0.0356 |
| *p<0.05, **p<0.01, ***p<0.001, ****p<0.0001 | | | | |
